# Information Theory-Guided Detection of Biomarkers Using Programmable Aptamer Arrays

**DOI:** 10.1101/2025.11.27.25341165

**Authors:** Amit Eshed, Alexander A. Green

## Abstract

While numerous endogenous nucleic acid biomarkers have been reported that are predictive for diseases like cancer, there has been limited development of devices that are tailored towards point-of-care (PoC) diagnosis using those endogenous biomarkers. Diagnosing based on endogenous biomarkers is challenging as its diagnostic value is based on concentration deviations from a healthy baseline. Here we report the Concentration Detector Array (CDA), a nucleic acid device that uses channel activated thresholds to identify and bin target nucleic acids into concentration ranges. Using consensus voting and error correction methods, we demonstrate that CDAs have excellent binning classification with an AUC = 0.945-0.955. Motivated by the diagnostic gap in non-small cell lung cancer detection, we developed probability distribution functions (PDFs) of prognostic miRNAs. After creating a library of miRNA sensing CDAs and PDF models, we demonstrated information theory-based diagnostic strategies to identify and classify patient profiles in a minimal number of tests. Our divergence maximization strategy was found to strongly identify profiles in a single test and learn maximal information content 1-2 tests faster than other expectation maximization strategies and use significantly fewer tests static non-learning methods required.

## INTRODUCTION

RNA and DNA biomarkers have proven to be valuable diagnostic material for bacterial^1^, viral^2–4^, and mutational sensing^5,6^. Additionally, as precursor transcripts for proteins and catalytic RNAs, nucleic acids are convenient markers for *in vivo* sensing of metabolic activity^7–9^. As a polymeric material, nucleic acids have incredibly useful features with their programmable base-pair binding rules, predictable secondary structures, and compatibility with a variety of fluorescent and colorimetric readout systems^7,10,11^. The universality of nucleic acids, and their fundamental characteristics have enabled the development of DNA and RNA devices for molecular detection, addressing various challenges in diagnostics and point-of-care (PoC) testing.

There have been numerous devices developed for nucleic acid sensing diagnostics based on technologies like molecular beacons, CRISPR-Cas, and toehold switches^2,12,13^. Recent advances in toehold-mediated strand-displacement platforms like the aptaswitch^14^ and FARSIGHT^15^ have demonstrated colorimetric signal readouts for PoC-aligned testing. However, the bulk of these nucleic acid detection methods have been focused on sequence identification of exogenous nucleic acids from pathogens. For many diseases, like some cancers, there may not exist a unique exogenous biomarker that is prognostic of illness. This necessitates the advancement of devices sensitive to endogenous biomarkers whose diagnostic value is based on deviations from a healthy concentration baseline, rather than simply its presence in a sample.

A variety of toehold-mediated strand-displacement platforms have been developed for concentration-dependent thresholding. Strand displacement of input/output targets from source gates has been applied in DNA neural network circuits that achieved pattern recognition and memory storage^16,17^. Concentration-dependent signaling systems have also been included in nucleic acid circuitry through logic gates, analog-to-digital converters, and diagnostic biosensors^13,18,19^. Other thresholding platforms have tuned the binding affinity between aptamers and proteins and protein-protein interactions in quantitative sensing and as bandpass filters in cellular circuits^20,21^. Many thresholding systems are challenging to apply for PoC platforms, with the reliance on multistrand systems requiring precise control over stoichiometry. The utilization of enzymes or modified nucleobases to control signal readout also increases assay cost and complexity.

Although molecular logic and thresholding systems have been applied towards diagnostics, the application of information theory to enhance diagnostic performance has not been demonstrated in these types of platforms. Information theory is typically used to solve optimization problems towards the transmission of information^22^. It has been previously applied in several clinical case studies, including characterizing the efficacy of alcoholism questionaries to reduce diagnostic uncertainty^23^, directing electrocardiograph and angiograph stress tests based on symptomatic presentations^24^, and evaluation of MRI imaging for prostate cancer pathology^25^. Additional demonstrations include comparing SARS-CoV-2 test diagnostic efficacy^25^ and for determining the criteria needed to rule-in/out disease states in diagnostic management plans for deep-vein thrombosis^26,27^. While these case studies illustrate the application of pre-existing technologies and assays for information theory-guided diagnoses, there has been little development of systems purpose-built for and capable of being optimized around an information-based diagnosis.

Appropriate uses of information theory for molecular diagnostic systems can enable significant decreases in assay complexity, increase confidence in results, and sharply reduce testing time. Information theory can guide the design of novel systems by developing an optimized symbol code for data compression, maximize channel capacity, and indicate the ordering of tests through expectation maximization. Moreover, it could direct better use of limited sample availability, for instance, a significant challenge in PoC diagnostic tests are limitations with patient samples volumes and clinical confidence such as peripheral blood or saliva over solid tissue biopsies. While accessibility to more complex biopsies are available in clinical settings, solid tissue sources do not necessarily provide an optimal route for diagnoses as biopsies can be slow, expensive, highly invasive, and often require multiple biopsies weeks to months apart for a clinically accurate diagnosis^26,28^. As such, principles from information theory can guide systems towards better resource optimization with less invasive samples to achieve efficient clinical diagnoses.

Here we report the Concentration Detector Array (CDA), a class of unimolecular aptamer-based devices that generate prescribed combinations of fluorescence signals when a nucleic acid biomarker falls within defined concentration ranges. The binning of the target biomarker based on concentration allows for the recognition of that biomarker from the healthy to disease states, providing the necessary context for distinguishing and monitoring endogenous biomarkers. Integration of all CDA sensing and output elements within a single RNA molecule avoids the need for precise stoichiometric control over different sensor elements and firmly links the signal characteristics of each output channel within a given CDA design. Moreover, use of fluorogenic aptamers within CDAs enables them to operate without requiring any modified bases. By creating a simplified circuit model of the CDA we analyzed how the features of individual devices influence their thresholding properties. Additionally, we created a library of devices for the detection of prognostic miRNA biomarkers for non-small cell lung cancer (NSCLC): miR-30a, miR-30d, miR-148a, and miR-182^29^. Lastly, inspired by information theory, the CDA devices were used to demonstrate a general diagnostic strategy applied to NSCLC. The successful application of information theory-based diagnostic strategies addresses a central challenge in PoC diagnostics, balancing the desire for optimized detection sensitivity while using the minimal number of tests. The outcome of these approaches can result in lessening the burden on patient sample acquisition, while learning as much as possible about the biomarkers necessary for classifying patient health status.

## RESULTS

### Concentration detector array design and mechanism

CDAs make use of previously developed aptaswitch devices^14^ that detect a cognate biomarker nucleic acid to activate folding of a reporter aptamer (**Fig. 1a**). Each CDA is composed of multiple aptaswitches encoded within a single transcript, wherein each aptaswitch has a unique fluorescent reporter aptamer selected for readout orthogonality. Accordingly, each CDA contains multiple independent fluorescent readout channels. It is advantageous to have all channels physically on the same transcript, as the channel ratios are intrinsically fixed to the design of the CDA. Moreover, a consistent 1:1 stoichiometry between aptaswitch channels is maintained, eliminating variability based on sensor copy number that can occur in multimolecular quantitative platforms. Each aptaswitch channel in the CDA has an occluding hairpin, which contains in the descending stem a **B\*** domain whose release is necessary to bind to a **B** domain downstream of the channel’s respective aptamer. When **B\*** and **B** hybridize, the core aptamer sequence between these domains is stabilized, inducing fluorescence. Each CDA channel is responsive to the same biomarker, but with varying sensitivities based on the length of the single-stranded toehold domain upstream of the occluding hairpin. The toehold domain serves as the initial binding domain of a biomarker for its respective channel. The activation mechanism for each channel occurs through toehold-mediated strand-displacement. First, the biomarker binds to the toehold domain. The **B\*** and **C\*** domains from the biomarker then displace the **B\*** and **C\*** domains in the hairpin, consequently releasing the vital **B\*** from the hairpin to activate that channel’s aptamer.

**Fig. 1.**
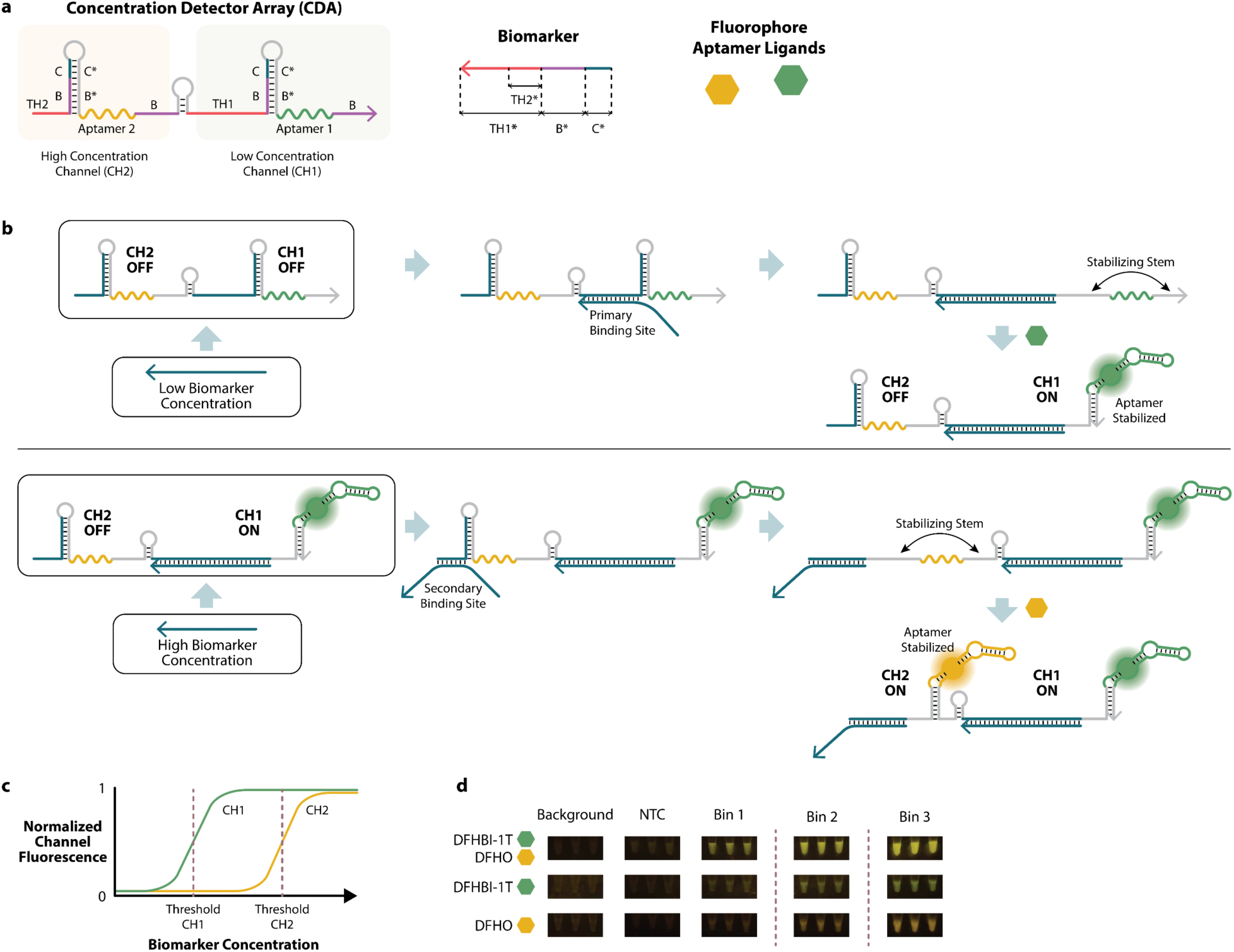
Design and activation mechanism of the concentration detector array (CDA). **a**, CDAs consist of multiple aptaswitches fused together in a single transcript and each designed to detect the same biomarker at different threshold concentrations. **b**, Binary channel activation mechanism of a two-channel CDA at low and high biomarker concentrations. The CH1 aptaswitch with the longer toehold domain activates at lower concentrations of biomarker, while the CH2 aptaswitch with the shorter toehold activates at higher concentrations. Channel activation stabilizes the reporter aptamer structure, enabling binding of the aptamer ligand to generate the readout signal. **c,** Expected emergent behavior of a CDA across biomarker concentration series with each output channel exhibiting a different activation threshold. **d**, Fluorescent readout on transilluminator after 10-minute incubation at 37°C. Top row with DFHBI-1T and DFHO fluorophore aptamer ligands present, middle row with only DFHBI-1T, and bottom row with only DFHO. NTC indicates a “no-target control”.

A low concentration channel, CH1, which activates at relatively lower concentrations of the biomarker nucleic acid, is identified as the channel with the longer toehold domain, **TH1** (**Fig. 1b**). A higher concentration channel will have a toehold domain, **TH2**, that is a shorter subsequence of **TH1**. When the CDA is exposed to increasing biomarker concentrations, CH1 will activate first, followed by the higher concentration channel when additional biomarker copies are available. On the population level, the CDA response to biomarker concentrations reaches signal equilibrium with exposure to higher concentrations of biomarkers equilibrating faster (**Supplementary Fig. 1**). **TH1**, as the longer toehold domain, serves as the CDA’s primary binding site for a biomarker. The more nucleotides that base-pair match to the biomarker the more favorable that channel is to be activated. The activation of the high concentration channel, CH2, intentionally does not maximize the full potential for base-pair binding to a biomarker and consequently makes **TH2** the secondary binding domain for the CDA device (**Fig. 1b**).

In a reaction with numerous CDAs and biomarkers available to hybridize, the binary activity of individual devices creates an emergent thresholding property as a population of CDAs becomes activated. The threshold property of a channel is used to define the population-level binary status that channel can be in, either ON or OFF denoted as a value of 1 or 0. In an idealized device, CH1 with the primary toehold **TH1** activates at a lower threshold concentration than CH2 with the shorter **TH2** (**Fig. 1c**). This thresholding property can be identified as early as 15 minutes into the device’s response to the concentration of an input biomarker (**Supplementary Fig. 2**). The design and mechanism of the CDA can be generalized and extended to an arbitrary number of channels, provided that each channel that activates at a higher concentration has a toehold domain that is shorter or has a lower melting temperature than the previous channel.

The number of bins a CDA can classify a biomarker into is one more than the number of channels it has. For a 2-channel CDA, the first bin is defined where both channels are OFF and ranges from a biomarker concentration of 0 µM to Threshold_CH1_. The second bin is defined as where CH1 is ON while CH2 is OFF and ranges from Threshold_CH1_ to Threshold_CH2_. The last bin, where both channels are ON extends from Threshold_CH2_ upwards. Devices were screened over a concentration series spanning 0.01 µM to 50 µM, to identify the channel thresholds and therefore in the analysis of devices, the practical upper bound limit of the third bin was set to 50 µM. These bins can be visually observed by fluorescent changes in channel activation due to sensitive biomarker concentrations. After only 10 minutes incubation at 37°C, the cognate fluorophore ligand, DFHBI-1T, for the low concentration channel is visually activated in bins 2 and 3, but not bin 1, while the cognate fluorophore ligand, DFHO, for the high concentration channel is distinctively activated in bin 3 and only moderately so in bin 2 in an example CDA. For both channels, fluorescent activation is minimal in bin 1. When both fluorophores are present, fluorescent activation per bin changes both in intensity and in hue (**Fig. 1d**).

### Two-channel and three-channel CDA activation

All experiments were prepared at consistent CDA and fluorophore concentrations and environmental conditions, with no-biomarker and background controls (see Methods). In a 2-channel CDA example, the difference in channel activity, with Broccoli aptamer^30^ in CH1 and Corn aptamer^31^ in CH2, was observed (**Fig. 2a**). CH1 saturated at a lower fold-change of around 9x while CH2 saturated with a fold change around 18x. The difference in fold change between channels may be due to several factors including the K_d_ between the aptamer and fluorophore, concentration and quantum yield of each fluorophore, the relative stability of each aptamer, and most significantly the leakiness of each channel when there is no biomarker present. The difference in channel thresholding was more readily observed in **Figure 2b,c** where we normalized each channel to the maximum fluorescence of the screening series. Each channel’s activation behavior was then fitted to a logistic regression function. Validated with high R^2^ values, channel activation was modeled as a logistic function since there are two distinctive plateaus in a channel’s activation signifying the OFF and ON states.

**Fig. 2.**
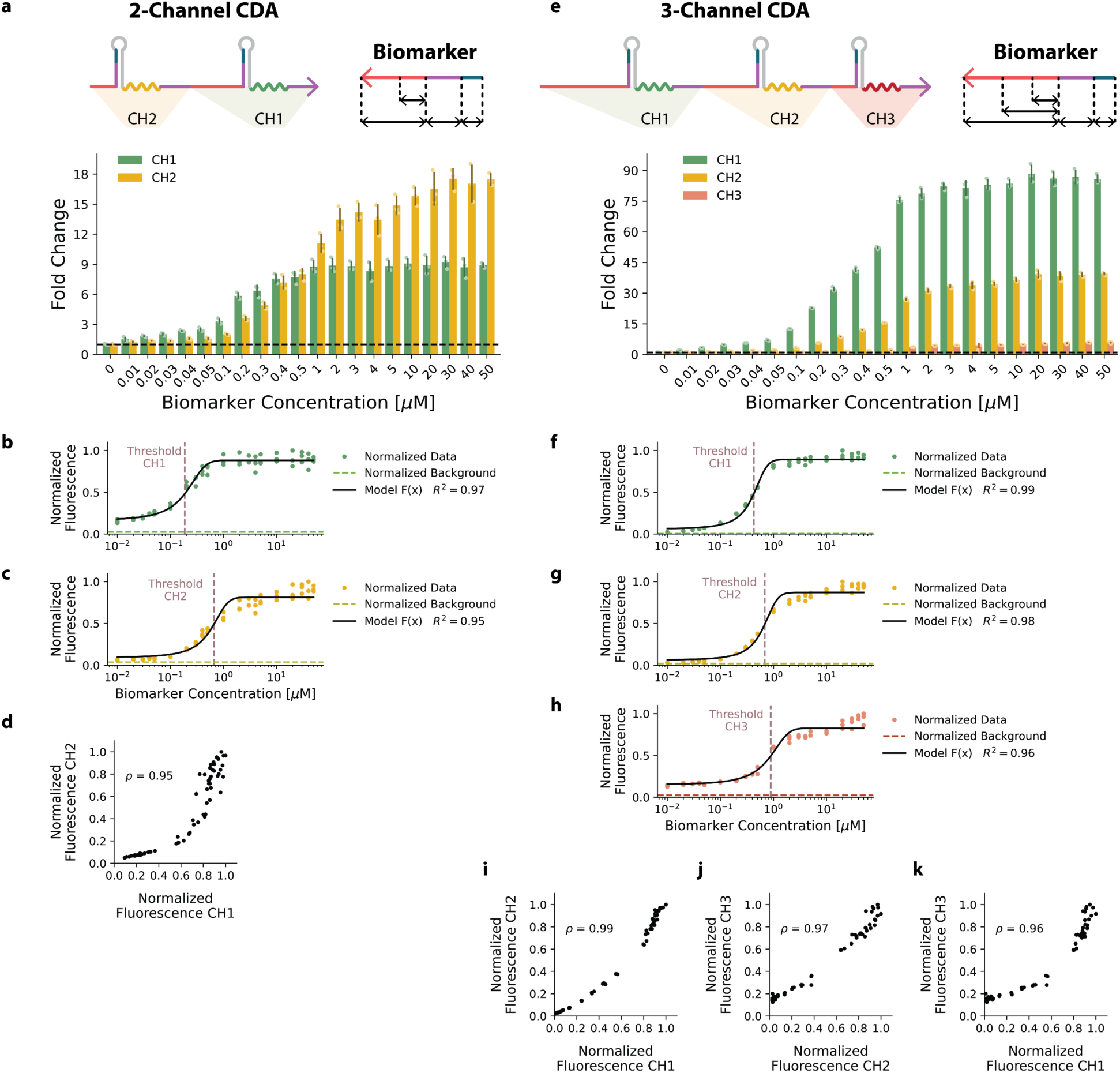
Two- and three-channel concentration detector arrays. **a**, Fold change of channel 1 and channel 2 fluorescence of a 2-channel CDA across a concentration series of biomarkers. Dashed line indicates a fold change of 1x. **b,** Channel 1 data normalized to the maximum fluorescence of the concentration series. Normalized data fitted to a logistic curve R^2^ = 0.97, Threshold_CH1_ = 0.186 µM. **c,** Channel 2 data normalized to the maximum fluorescence of the concentration series. Normalized data fitted to a logistic curve R^2^ = 0.96, Threshold_CH2_ = 0.663 µM. **d,** Normalized fluorescence of channel 2 in terms of channel 1. ρ = 0.95. **e,** Fold change of channel 1, channel 2, and channel 3 fluorescence of a 3-channel CDA across a concentration series of biomarkers. Dashed line indicates a fold change of 1x. **f,** Channel 1 data normalized to the maximum fluorescence of the concentration series. Normalized data fitted to a logistic curve R^2^ = 0.99, Threshold_CH1_ = 0.428 µM. **g,** Channel 2 data normalized to the maximum fluorescence of the concentration series. Normalized data fitted to a logistic curve R^2^ = 0.98, Threshold_CH2_ = 0.688 µM. **h,** Channel 3 data normalized to the maximum fluorescence of the concentration series. Normalized data fitted to a logistic curve R^2^ = 0.96, Threshold_CH3_ = 0.893 µM. **i,** Normalized fluorescence of channel 2 in terms of channel 1. ρ = 0.99. **j,** Normalized fluorescence of channel 3 in terms of channel 1. ρ = 0.96. **k,** Normalized fluorescence of channel 3 in terms of channel 2. ρ = 0.97. N = 3, error bars indicate ±1 standard deviation from the mean. R^2^ indicates the coefficient of determination correlation. ρ indicates Spearman rank correlation coefficient. Black dashed line in figures **a** and **e** indicates a fold change of 1. Normalized background for **b**, **c**, **f**, **g**, and **h** indicates the ratio fluorescence of the background control to the maximum fluorescence of the concentration series. Solid black line in **b**, **c**, **f**, **g**, and **h** indicates least-squares fitted logistic model. Maroon dashed lines in **b**, **c**, **f**, **g**, and **h** indicate the threshold concentration as the inflection point of the logistic model.

The thresholding concentration for each channel, as designated by the maroon dashed line, was calculated as the biomarker concentration at the inflection point of the logistic model. Here we calculated the thresholding concentration for CH1 and CH2 as 0.186 µM and 0.663 µM respectively. To understand the relative channel behaviors of the CDA, we plotted percent activation of CH2 in terms of CH1. In an idealized device, CH1 should activate completely before CH2 starts to turn-on. We observed with this CDA, that CH1 activated from 0% to approximately 70% with about 30% leakage in CH2 (**Fig. 2d**). For all CDAs we also observed, regardless of biomarker and domain lengths, a negative exponential relationship between the concentration threshold of a channel and its local derivative from the fitting model (**Supplementary Figure 3**). The threshold-derivative relationship suggests that channels that switch on at lower concentrations are also more sensitive to concentration changes than channels that activate at higher concentrations.

The CDA design was also extended to a 3-channel array, subdividing a concentration series into 4 bins between 0 µM to Threshold_CH1_, Threshold_CH1_ to Threshold_CH2_, Threshold_CH2_ to Threshold_CH3_, and Threshold_CH3_ to 50 µM. In a 3-channel CDA, CH1 represented by Broccoli had the highest fold change, saturating at around 85x. CH2 represented by Corn and CH3 under Mango-IV aptamer^32^ each saturated at a lower fold change of approximately 40x and 6x respectively (**Fig. 2e**). Following a similar analysis as the 2-channel arrays, we calculated the thresholding concentrations for CH1, CH2, and CH3 to be 0.428 µM, 0.688 µM, and 0.893 µM, respectively (**Fig. 2f-h**). When directly comparing channel activation behaviors for the 3-channel CDA, we again observe similar concavity as the 2-channel CDA (**Fig. 2i-k**) as well as a decreasing derivative rate at higher thresholds.

### CDA characterization and functionality

In the process of developing a library of functional 2-channel CDAs we varied several design parameters. Across several iterations, we perturbed the lengths of the four key domains (**TH1**, **TH2**, **B**, and **C**) that describe individual CDAs and channels, as well as the total length of the biomarker, the ordering of channels along the RNA strand, which aptamer represents which channel, and the inclusion of a spacer or hairpin motif to form a barrier between channels. For the 62 devices first developed and screened, we calculated Threshold_CH1_ and Threshold_CH2_ and found that 31 were deemed functional. We labeled devices as functional if they followed the conceptual inequalities, that being, CH2 with the short **TH2** activates at a higher threshold than CH1 with the long **TH1**.

Devices that were labeled as nonfunctional may not have worked as intended for a few reasons. One condition for a device labeled nonfunctional was if the inequality relationship did not hold true, that CH2 was activated at a lower threshold than CH1. Another condition was if signal activation was invariant to the concentration of the biomarker. This behavior may indicate one of two errors, the device does not turn on in the presence of its biomarker, observable by fluorescence of experimental groups being indistinguishable from background levels. Alternatively, the channels were too leaky, as indicated by a high fluorescence from the no-target control and consequently adding biomarkers would not increase channel signal any further.

To identify design rules for CDAs, we first explored how individual design features influence functionality. The design features investigated included the domain lengths of **TH1**, **TH2**, **B**, and **C** that describe CDAs, as well as Threshold_CH1_ and Threshold_CH2_ (**Supplementary Fig. 4**).

Across variations in the lengths of the domains **TH1**, **TH2**, **B**, and **C**, there was a poor distinction between the domain lengths of functional and nonfunctional devices as designated by a weak regression fit classifier. The **C** domain, which was the best domain length classifier, achieved a device functionality classification sensitivity of 84%, specificity of 36%, and accuracy of 60%, but had a regression fit R^2^ of only 0.13. Threshold_CH1_ had a weak functionality classifier with an R^2^ of 0.21. Threshold_CH2_ had a moderately strong regression fit to explain functionality with an R^2^ of 0.74 and a classification sensitivity, specificity, and accuracy of 94%, 87%, and 90% respectively.

From **Supplementary Fig. 4**, we can draw a general conclusion that variations in **TH1**, **TH2**, **B**, **C**, and Threshold_CH1_ features do not explicitly explain why some of devices worked and others did not. Variations in Threshold_CH2_ might suggest that there may be a minimum threshold concentration, approximately 290 nM at the inflection of the classifier, for which devices are functional.

Since the relationships between **TH1** and **TH2**, and Threshold_CH1_ and Threshold_CH2_ are innate to how we defined functionality, we considered whether functionality of devices can be explained as a relationship of multiple features simultaneously. We conducted a PCA analysis with the four individual design features (**TH1**, **TH2**, **B**, and **C**) and the two emergent features Threshold_CH1_ and Threshold_CH2_. Interestingly, when we ran a PCA analysis of the six features and labeled the devices with their functionality, we found a distinctive separation between functional and nonfunctional devices along principal components 1 and 2 with a combined explained variance of 82.3% (**Fig. 3a**). As shown in **Supplementary Figure 5**, there is also some distinction between functionality when comparing principal components 1 to 3 and 2 to 3. We next fitted a linear decision boundary to separate functional and nonfunctional devices in **Fig. 3a**, defining functionality regions in PCA space. The decision boundary line had a functionality classification sensitivity, specificity, and accuracy of 90.3%, 74.2%, and 82.3%, respectively. To test the robustness of the decision line, we made a new boundary fit with a randomly selected training set of 10 functional and 10 nonfunctional devices and remeasured the classification capabilities with the remaining 42 labeled devices (**Supplementary Fig. 6**).

**Fig. 3.**
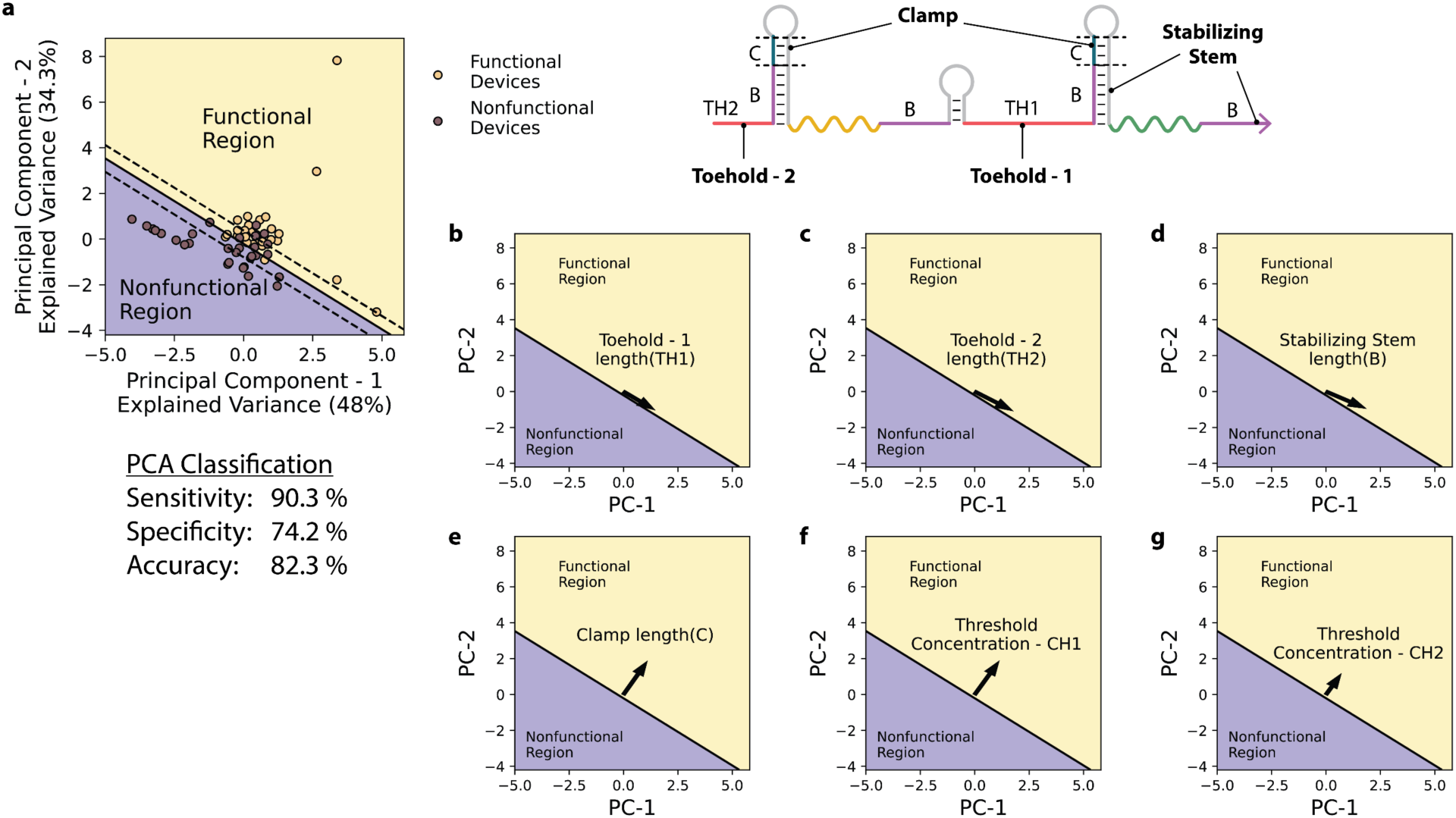
Principal component analysis of concentration detector array features indicate functionality. **a**, PCA classification of functional and nonfunctional devices. Solid line indicates the 0-contour line of the decision boundary curve; the upper and lower dashed lines indicate the 0.95 and −0.95 contour lines of the decision boundary curve between classifying functional devices labeled as value 1 and nonfunctional devices labeled with value of −1. **b-e,** Loading scores of PCA feature lengths of **TH1** (**b**), **TH2** (**c**), **B** (**d**), **C** (**e**) plotted as a vector. **f-g,** Loading score of PCA feature Threshold_CH1_ (**f**) and Threshold_CH2_ (**g**) plotted as a vector. All loading score magnitudes are multiplied by a factor of 2 for better visualization on PCA vector plots **b-g**.

To gain insight into why there exists a distinction between functional and nonfunctional devices in PCA space, we plotted the loading scores of the PCA features (**Supplementary Table 1**) as vectors. As seen in **Fig. 3b-g**, we found that three features (**TH1**, **TH2**, and **B**) appear to be nearly parallel to the decision boundary line, while three features (**C**, Threshold_CH1_, and Threshold_CH2_) appear to be nearly orthogonal to the line. The angles between the **TH1** vector and **TH2** vector to the boundary line were 8.4° and 12.2° respectively. With a greater angle from the boundary, length increases in **TH2** may support CDA functionality to a greater extent than **TH1**, although both features have acute angles. As recalled in **Supplementary Fig. 4**, the **B** domain showed a weak functionality classification, which is supported by its small vector angle from the boundary line at 16.6° (**Fig. 3d**).

The **C** vector angle was calculated at 103.3° which may provide an insight into why some devices failed (**Fig. 3e**). The function of the clamp, **C** domain, is to make the channel OFF state energetically favorable in the absence of the biomarker. If the clamp domain is too short, the occluding hairpin may not be strong enough to consistently sequester the necessary **B\*** domain from stabilizing the aptamer. Fluorescence in the absence of a biomarker will make channel activity less sensitive to the presence of the biomarker and consequently affect the calculated threshold. An increase in the length of the clamp domain would reduce channel leakage. Based on its large angle from the boundary line, increasing the clamp domain may be the most direct way to improve the likelihood of a CDA being functional through its design.

Threshold_CH1_ (**Fig. 3f**) and Threshold_CH2_ (**Fig. 3g**) were both calculated to be 102.8° from the boundary line. Interestingly, while the Threshold_CH1_ and Threshold_CH2_ vectors are parallel to each other, their magnitudes are different, with a unit increase in Threshold_CH1_ moving a device in PCA space about 2.5x further into the functional region than Threshold_CH2_. While changes in thresholding concentration for both channels will move a device in PCA space in the same direction, the difference in magnitudes suggests some limitations about the relationship between Threshold_CH1_ and Threshold_CH2_. For a device to be classified as functional, Threshold_CH1_ must be smaller than Threshold_CH2_. However, in PCA space, an increase Threshold_CH1_ relative to Threshold_CH2_ suggests that a device is more likely to be functional. These two opposing ideas about functionality can be understood as a practical limitation deciding how distinct each channel threshold can be from the other.

### Operational amplifier comparator circuit model

With a better understanding of what made devices functional, we next sought to create a model of how the design features (**TH1**, **TH2**, **B**, and **C**) may influence the emergent thresholding properties of the CDAs. As such, we modeled the CDA devices as an op-amp comparator circuit (**Fig. 4a**). The governing equations for the model create a circuit (see **Supplementary Note 1**) whose V_OUT_ in terms of a V_IN_ voltage sweep resembles the channel activation experimentally observed by the CDAs (**Fig. 4b**).

**Fig. 4.**
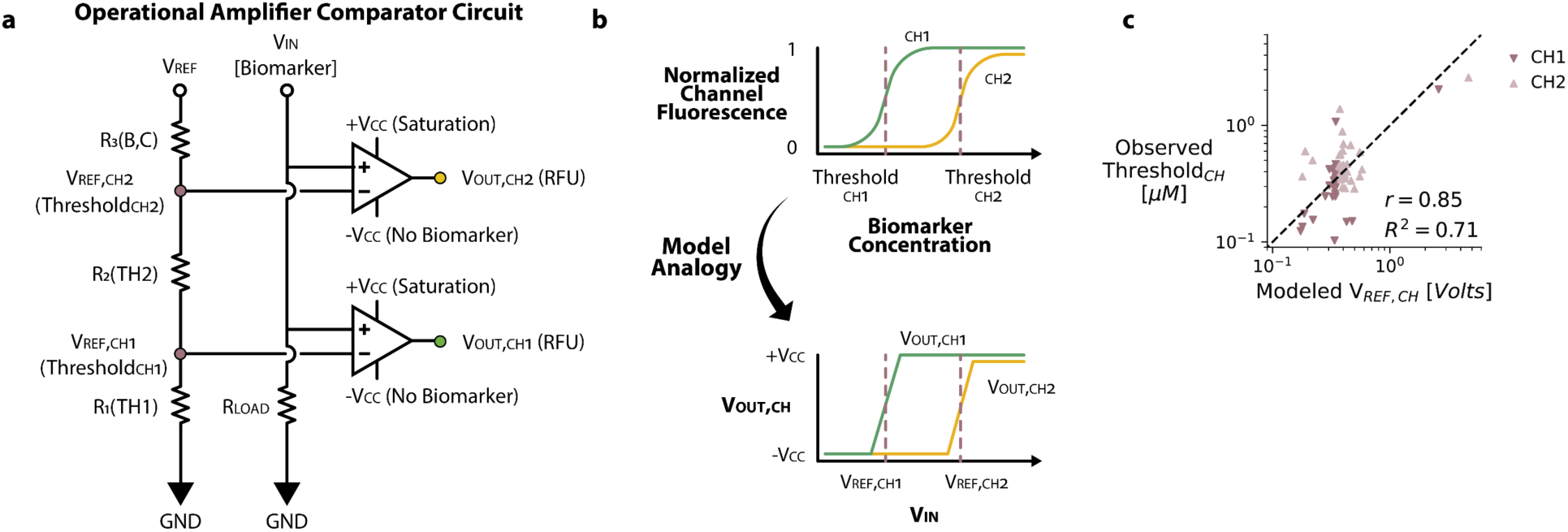
Operational amplifier comparator circuit model of concentration detector arrays. **a**, Operational amplifier comparator circuit model of CDA devices. **b,** Model analogy expected fluorescence output of CDA devices behaves similarly to op-amp comparator circuit. **c,** Comparison of modeled voltage thresholds versus experimental observed threshold concentrations. Pearson correlation coefficient r = 0.85, coefficient of determination R^2^ = 0.71.

To connect this circuit analogy to the design features of the CDA, we set the resistors R_1_, R_2_, and R_3_ as a function of the domain lengths **TH1**, **TH2**, and **B** and **C**, respectively as these are the domains that must interact with the biomarker to initiate toehold-mediated strand-displacement and activate the fluorescent aptamer. It was determined that **TH1**, **TH2**, and **B** domain lengths were set to be inversely proportional to the resistance as these domains are influential in moving the strand-displacement process forward. From the OFF to the ON state, there is a net increase in base-pairs formed by these domains making the forward kinetic reaction favorable. Analogously, a net increase in base-pair binding acts like a conductance, enabling a more energetically favorable folding state of the device, providing less “resistance” towards channel activation. The **C** domain was set to be proportional to resistance. As previously described, the clamp domain aids in sequestering the **B\*** domain in the hairpin resisting channel activation. Additionally, in the presence of the biomarker, there is no net increase in base-pair hybridization of the **C** domains, as the **C\*** in the hairpin is displaced by the **C\*** from the biomarker.

The op-amp comparator model was able to predict V_REF,CH1_ and V_REF,CH2_ to the observed Threshold_CH1_ and Threshold_CH2_ of the 31 functional devices (62 data points), achieving a Pearson correlation of 0.85 and R^2^ of 0.71 (**Fig. 4c**). While this model is moderately strong, we consider it in its context as an analogy to learn from and help design new CDAs. Unexplained variability in the correlations can be attributed to numerous other features that were not included in the model such as the total length of the biomarker, the GC content of each domain, the 5’ or 3’ ordering of each channel, and which aptamers represent which channel. A more detailed model could be created by considering additional variables; however, this would require a much larger dataset of functional devices to prevent the risk of overfitting.

### CDA binning and recognition capabilities of non-small cell lung cancer (NSCLC)-predictive miRNAs

Lung cancer is the second most common cancer type for both men and women and has the greatest number of deaths by cancer type^33,34^, which is attributed to it being undiagnosed until it is in an advanced stage with over 75% of patients being diagnosed at stage III or IV^35,36^. The delay in diagnosis often leads to avoidable cancer related deaths, as the five-year survival rate for patients diagnosed with stage I lung cancer is 68.4%, but drops significantly to 26.2% at stage IIIA, and down to only 5.8% when diagnosed at stage IV^35,36^.

To address the diagnostic gap in lung cancer detection, highly accessible PoC-aligned platforms need to be developed. The most common imaging method for diagnosis in lung cancer screenings, low dose computed tomography (CT), has low utilization with approximately 18% of age and risk recommended patients getting screened^37^. The most invasive but clinically significant testing, involving solid tissue biopsies, can be used to identify patient tissue samples for characteristic phenotypes of cancerous cells through immunohistochemistry labeling of identifier proteins and biomarkers^38^. CT scans and solid tissue biopsies however, are unfavorable for PoC-testing due to the expensive instruments, trained personnel required, and complex materials that need to be analyzed, adding weeks to months to diagnose before treatments can begin^28,35,38^. Blood chemistry tests used to profile prognostic biomarkers can identify the dysregulation or mutagenesis in patient samples indicating deviations from normal cellular proliferation rates^35,38^. While molecular detection assays that target prognostic biomarkers might not provide the same clinical strength to diagnose as other methods, these assays can serve a vital role in diagnosis for quickly and efficiently determining disease risk in PoC settings and then direct a patient to seek clinical confirmation if high risk^35^.

miRNAs are a promising biomarker for cancer diagnostics due to their high stability and differential expression of healthy from disease concentrations in minimally invasive blood samples, even at stage I lung cancer cases^35,36^. While the expression profiles of miRNAs promise a strong predictive power, there are some general challenges to detecting miRNAs. miRNAs, as regulatory markers, characteristically become dysregulated in cancers, as certain miRNAs are often found to be correlated with several cancer types^39–41^. Additionally, between cancer types and stages, miRNAs can be expressed over or under their baseline healthy concentration ranges necessitating detection over a range of potential miRNA concentrations^10,36,40^. When creating a classifier model for NSCLC, Zhang et al., identified a combined expression profiles of miR-30a, - 30d, −148a, and −182 to have a significant labeling accuracy for tumor and healthy samples^29^. As such, we created a new library of 2-channel CDAs for the detection of these prognostic miRNAs for NSCLC. After validating a library of miRNA targeting CDAs, we sought to characterize the efficacy of CDAs towards successful identification of a target biomarker into bins.

A CDA can locate a target biomarker within defined bins by its thresholds (**Fig. 5a**). When an unknown profile is tested against the CDA, the output fluorescence from both CH1 and CH2 are measured and normalized to each channel’s respective maximum fluorescence as previously characterized. Each channel will “vote”, indicating if the normalized fluorescence of the profile is above or below the fluorescence at that channel’s threshold concentration. With each test, both CH1 and CH2 will fluoresce to a certain percent activation and simultaneously indicate which bins each threshold identifies the biomarker could be in. The output bin identified by the CDA is determined by which bin has consensus or greatest number of votes. If the CDA assigns a profile to the correct bin, then that bin is labeled as a true positive, and the other bins as true negatives (**Supplementary Fig. 7**). If the CDA makes a classification error, then the misidentified bin is labeled as a false positive, the correct bin as false negative, and the third bin as a true negative. As more CDAs are applied in parallel tests, the combined output from multiple devices allows for an improved consensus and convergence towards a bin with a smaller concentration range (**Fig. 5b**).

**Fig. 5.**
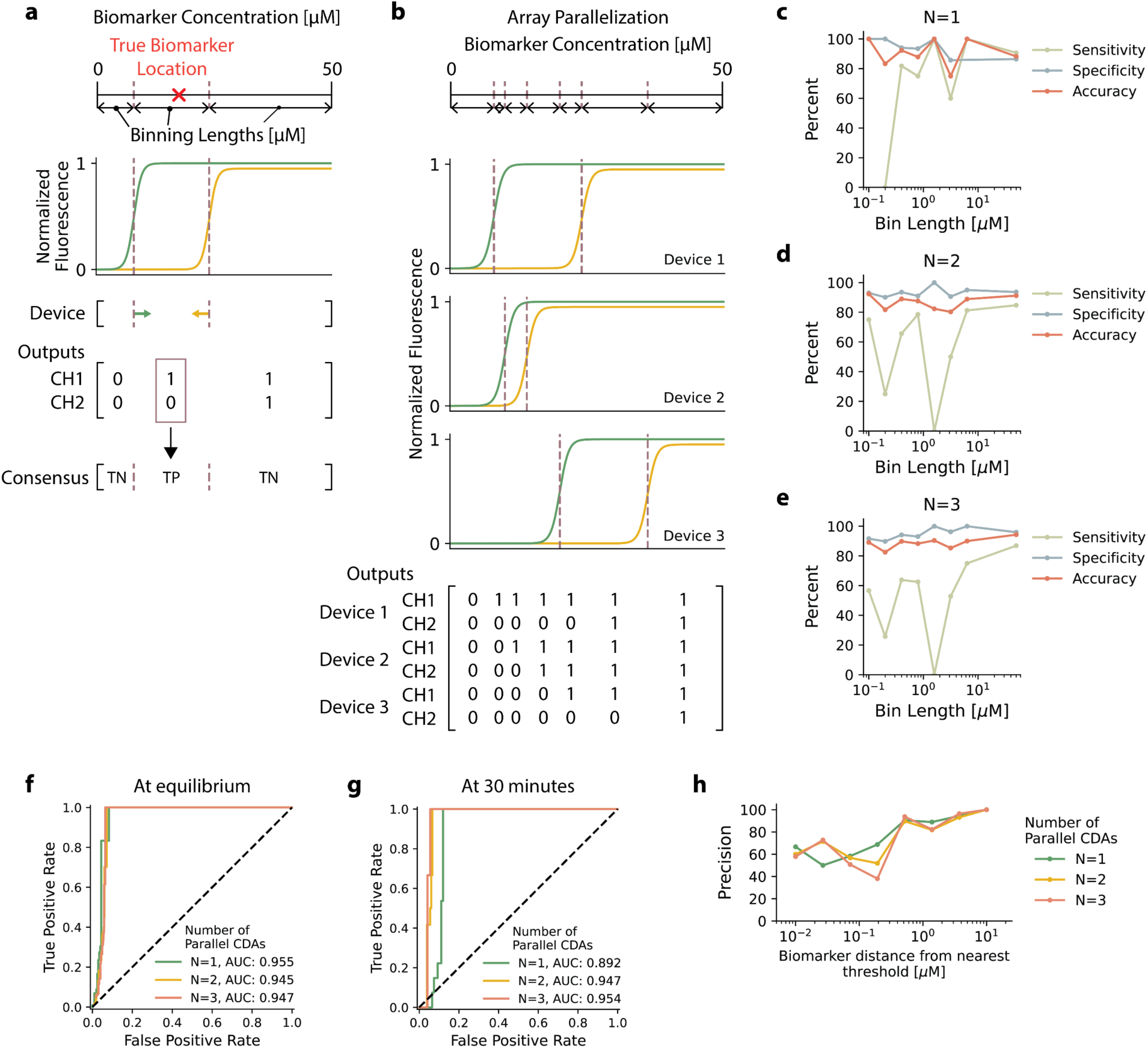
Concentration detector array parallelization and binning of prognostic biomarkers for non-small cell lung cancer. **a**, CDA bins a biomarker concentration based on the device’s innate thresholding concentrations. Binning classification is identified by the output activation of fluorescent channels. Binning length measures the width of a bin in µM. **b**, Application of several CDAs in parallel creates numerous bins. The joint information of all fluorescent outputs is used to classify a target into a particular bin. **c**, Bin classification sensitivity, specificity, and accuracy of library of CDAs targeting miR-30a, miR-30d, miR-148a, and miR-182 as a function of bin length. **d**, Bin classification sensitivity, specificity, and accuracy of library of two parallel CDAs targeting miR-30a, miR-30d, miR-148a, and miR-182 as a function of bin length. **e**, Binning classification sensitivity, specificity, and accuracy of library of three parallel CDAs targeting miR-30a, miR-30d, miR-148a, and miR-182 as a function of bin length. **f**, Receiver operator curve of binning classification of one, two, and three parallel CDAs when all device channels reach equilibrium, analysis at 6-hour timepoint. **g**, Receiver operator curve of binning classification of one, two, and three parallel CDAs for rapid testing, analysis at 30-minute timepoint. **h**, Binning classification precision of CDAs as a function of target biomarker distance from nearest threshold. CDA’s precision represented with one, two parallel, and three parallel CDAs.

Channel errors can also be discovered and corrected through consensus voting. If no errors are made, then there should only be one bin that has majority consensus. However, if one channel has a flipped directionality, then consensus may be split between two competing bins. In this circumstance, consensus is reestablished by creating the smallest continuous bin that merges the two disputed bins as illustrated in **Supplementary Figure 7**.

To validate the CDA binning capabilities, we challenged each device against six concentration values set to specifically fall in-between CDA thresholds. Per each test and bin of a CDA, we recorded the bin’s length in µM and the classification status as true positive, true negative, false positive, or false negative. We then calculated sensitivity, specificity, and accuracy of CDAs as a function of the bin lengths for one, two, or three parallel devices allowing for a greater resolution of binning lengths with more devices (**Fig. 5c-e**). While specificity and accuracy for bin classification seems to be consistent across bin lengths at 80-100%, regardless of the number of parallel devices, sensitivity is more variable. We also performed this same analysis per target miRNA and identified that there may be some sequence dependency as miR-30d and miR-148a targeting CDAs are more consistent across bin lengths than miR-30a and miR-182 CDAs (**Supplementary Fig. 8**). Despite the variable sensitivity, the CDA ability to effectively classify biomarkers into bins was quite robust with an AUC ranging from 0.945-0.955 (**Fig. 5f**). For rapid testing in 30 minutes, CDA bin classification is effective with one device testing AUC of 0.892. With the benefit of consensus voting and error correction through parallel tests, the AUC with two and three CDAs increases to 0.947-0.954 (**Fig. 5g**).

To better understand the source of variability in binning sensitivity observed in **Fig. 5c-e**, we mapped binning precision as a function of the target biomarker distance from the nearest threshold. As seen in **Fig. 5h**, we observed a decrease in precision as the biomarker is expressed closer to the threshold value, becoming a nearly random classifier at close distances to the threshold. As the biomarker approaches the decision-making threshold value, small fluctuations in noise and fluorescence signal are much more impactful, potentially causing a channel to flip directionality and vote towards an incorrect bin.

### CDA diagnosis of non-small cell lung cancer using information theory-based strategies

While binning resolution can be improved by testing more CDAs, there is a need to be economical for PoC testing and balance the number of tests that can be used on a patient sample against diagnostic confidence. In clinical settings, there are limitations on how much sample can be collected and utilized as biopsies are slow, expensive, and invasive^28^. For PoC settings, adoption of tests depends upon non-invasive and minimal sample volume acquisition. These challenges come against another limitation in diagnostic settings, that multiple tests may be required to establish a clinically accurate diagnosis^26^. Therefore, in diagnostic testing, particularly for PoC-aligned platforms, each test performed on a patient sample must provide as much useful information towards diagnosis as possible, ensuring that we make the best use of limited volumes and restrictive samples^25^. To establish an efficient diagnosis strategy, we used core concepts from information theory to optimize and maximize the amount of information learned about a biomarker while using as few tests as possible.

Information theory has historically been used to address problems in communications, data compression, and error correction based on probability distribution functions (PDFs) of certain states or symbol codes^22,25^. Here we applied information theory to characterize the diagnostic efficiency and efficacy of CDAs for NSCLC using information content, Shannon entropy (H), and the Kullback-Leibler divergence (D) (see **Supplementary Note 2**). Resolving an unknown random variable, like the concentration of a miRNA, into a certain concentration range requires learning information about the biomarker. Information content, measured in bits, quantitatively represents that amount of information learned once a bin is identified by a CDA test. It is based on the associated probabilities of the health state (healthy or tumor) integrated over that bin. As the probability of identifying a biomarker within a diminishing concentration range decreases, the amount of information learned increases. The Shannon entropy H represents the expected amount of information to be learned from a particular test. Here we use entropy as a pre-test measurement to characterize devices, describing how much information we expect to learn about a biomarker when using a particular CDA and assuming a specific PDF model. Kullback-Leibler divergence D describes the relative difference between PDF_Healthy_ and PDF_Tumor_ models when discretized by the thresholds of the CDA. For our application, divergence is used to characterize CDA’s classification efficiency. A device with a high divergence bit value will be more capable of distinguishing a patient profile of being associated with the PDF_Healthy_ from the PDF_Tumor_ model. With the characterized library of CDAs, we implemented several information theory-based diagnostic strategies to enable effective classification of profiles as healthy or cancerous in a minimal number of tests.

Based on the four prognostic miRNAs for NSCLC, we created a custom cohort (**Supplementary Table 3**) from The Cancer Genome Atlas (TCGA) database, collected all open access miRNA read counts from which to build our PDF models. The custom cohort was based on 739 peripheral blood aliquots from lung adenocarcinoma and lung squamous cell carcinoma patients, the two predominant cancer subtypes of NSCLC^42^, and 64 healthy samples. We first identified miR-1296 as a normalization marker due to it having an invariant expression regardless of health status (**Supplementary Fig. 9**). For each prognostic miRNA, we plotted a histogram of the tumor and healthy states using normalized read counts and fitted them to Gaussian PDFs (**Supplementary Fig. 10**).

Due to miRNA concentrations in patient samples being expressed below the thresholding sensitivity of CDAs, miRNAs needed to be amplified such that their DNA amplicons were expressed at the effective concentration ranges of the CDAs. To determine the optimal concentration that the miRNA PDFs would need to be amplified, we calculated the Shannon entropy of our entire CDA library as we shifted the means of the PDFs and identified at what concentration entropy was maximized (**Supplementary Fig. 11** and **12**). The PDFs for each miRNA and health state were shifted by the same amplification factor to the optimized concentration for the CDA library. Joint amplicon PDFs (**Supplementary Fig. 13**) were developed by cross multiplying the one-dimensional PDFs identified in **Supplementary Fig. 12**. With amplicon PDFs numerically established at the effective concentration range for our library of CDAs, we then detected amplified miRNAs and binned them according to the new PDF models (**Fig. 6a**).

**Fig. 6.**
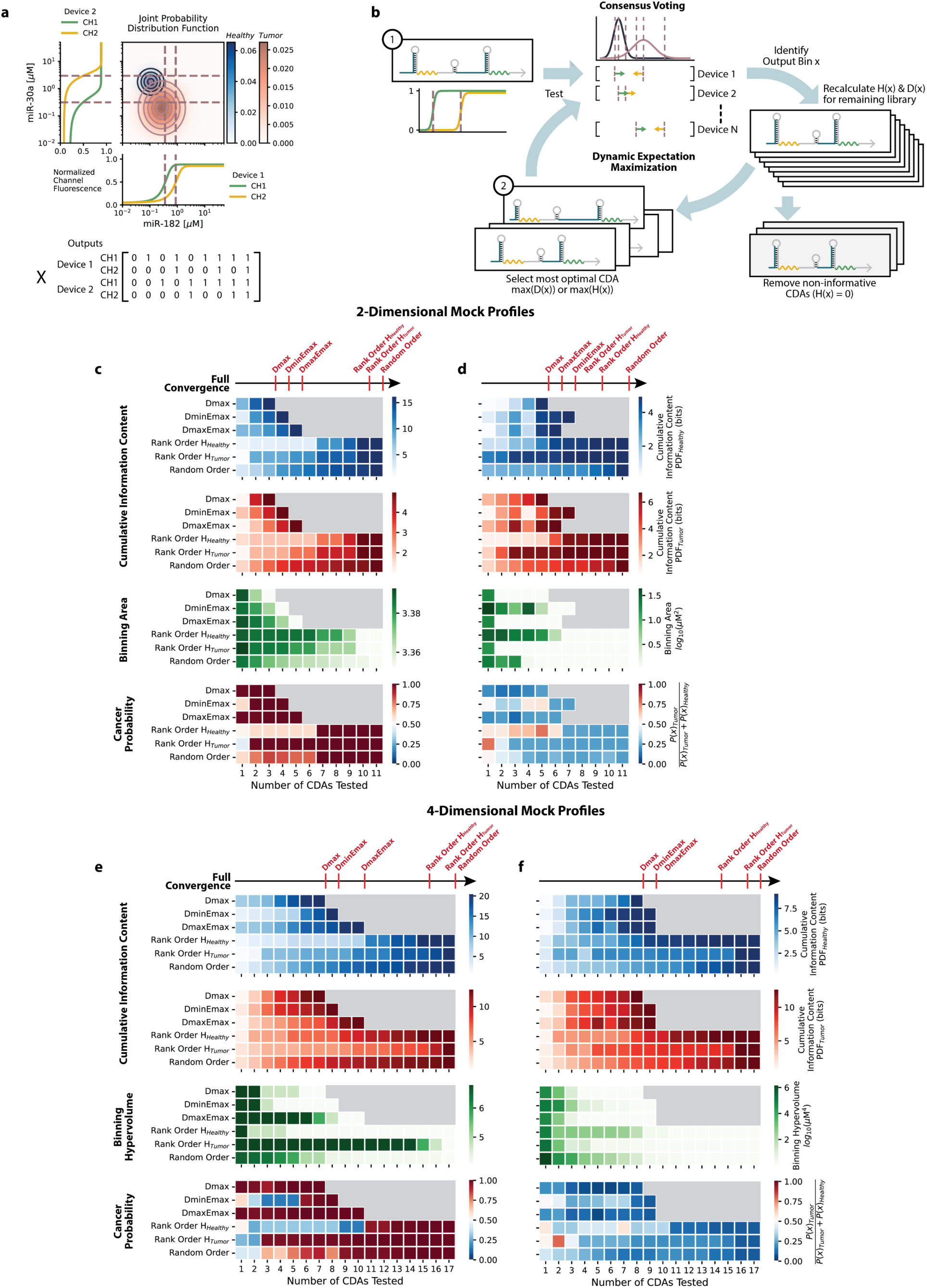
Information theory inspired diagnostic strategies to diagnose non-small cell lung cancer. **a**, Healthy and tumor joint probability distributions for miR-30d and miR-182 binned by two orthogonal CDAs identify nine potential binning outputs. **b**, Generalized process for dynamic diagnostic strategies. **c**, Diagnostic strategies demonstrate learning information assuming healthy PDF model, tumor PDF model, convergence on binning area, and classification of a mock tumor profile using a 2-dimensional PDF based on miR-30d and miR-182. **d**, Diagnostic strategies demonstrate learning information assuming healthy PDF model, tumor PDF model, convergence on binning area, and classification of a mock healthy profile using a 2-dimensional PDF based on miR-30d and miR-182. **e**, Diagnostic strategies demonstrate learning information assuming healthy PDF model, tumor PDF model, convergence on binning hypervolume, and classification of a mock tumor profile using a 4-dimensional PDF based on miR-30a, miR-30d, miR-148a, and miR-182. **f**, Diagnostic strategies demonstrate learning information assuming healthy PDF model, tumor PDF model, convergence on binning hypervolume, and classification of a mock healthy profile using a 4-dimensional PDF based on miR-30a, miR-30d, miR-148a, and miR-182. **c-f**, Full Convergence indicates per diagnostic strategy, the test at which the maximum resolution of the biomarker concentration range is achieved given a library of CDAs.

For each device, we calculated the entropy assuming the amplified PDF_Healthy_ and PDF_Tumor_ models, as well as the divergence (**Supplementary Fig. 14**). From **Supplementary Fig. 14**, we observed that several CDA devices approach the theoretical maximum entropic distribution of 1.58 bits. Using entropy and divergence we devised six diagnostic strategies, three static and three dynamic methods, that decide the most optimal order of CDAs to use for the diagnosis of patient profiles with the fewest tests (**Fig. 6b**). To monitor the progress of diagnosing a patient profile per test, we calculated the cumulative information content associated with each health status, the binning area, and the probability of a patient status being healthy or NSCLC.

Of the three static strategies, the first was a random order strategy, where devices were randomly selected without replacement from the library ten times, and the information content, binning areas, and probabilities were averaged over the ten repeats. The other two static methods are Rank Order H_Healthy_, and Rank Order H_Tumor_ (**Supplementary Fig. 15**). The rank order strategies test CDAs in order of decreasing entropy, as reported in **Supplementary Figure 14**, assuming either the PDF_Healthy_ or PDF_Tumor_ models respectively. By testing the most entropic devices first, it is expected that a CDA with a high H_Healthy_ will learn a lot of information about a profile if a patient is healthy, similarly a CDA with a high H_Tumor_ will be better optimized for NSCLC patients.

We also investigated three dynamic strategies that rely on the cumulative binning outcomes from the previous tests to inform which device from the remaining library should be used next (**Supplementary Fig. 16**). These strategies represent various forms of expectation maximization. Dmax, applies the CDA that has the greatest calculated divergence given the combined binning outputs from the previous tests. DminEmax’s first test uses the CDA that minimizes divergence, and then all subsequent tests using CDAs that maximize the entropy of the most probable health state. DmaxEmax’s first test uses the CDA that maximizes divergence, like the Dmax, and all subsequent tests maximize the entropy of the most probable health state. As dynamic strategies, it is expected that by using the information learned from previous tests, that these strategies will learn faster, converge rapidly onto the binning areas of the patient profile biomarkers, and classify the patient faster than the static non-learning methods. It is expected that dynamic entropic-based methods will learn faster than the dynamic divergence-based method. However, the Dmax is expected to differentiate and classify profiles in fewer tests between the healthy and tumor PDF models which is most realistic for our goals of developing a PoC-aligned diagnostic strategy.

To demonstrate the effectiveness of the diagnostic strategies, mock tumor and healthy patient profiles were created and tested. Profiles were developed by first randomly selecting patient profiles from the custom TCGA cohort, identifying the patient health state, and normalizing the read counts for the prognostic miRNAs to miR-1296. Based on the normalized read count PDFs created in **Supplementary Fig. 10**, the percentile for each miRNA from the patient profile was calculated. Using the amplified concentration PDFs created in **Supplementary Fig. 12**, the miRNA “amplicon” concentration was determined under the same percentile as the normalized read count PDFs. CDAs were tested with the mock patient miRNA “DNA amplicons” (DNA oligos of the target miRNAs) at the calculated concentrations.

A mock tumor and healthy profile based on miR-30d and miR-182 amplicons and the corresponding 2-dimensional joint PDF models from **Supplementary Fig. 13** were evaluated with the six diagnostic strategies (**Fig. 6c,d**). The three static methods generally learned the slowest, yielding the fewest bits per test. As seen in **Fig. 6c**, Dmax learned the maximum amount of information in only three tests and interestingly converged on the binning area the fastest, followed by the entropy-based maximization strategies. On the first test with a highly divergent CDA, both Dmax and DmaxEmax had high confidence in identifying the mock profile as tumor. We also observed the consequence of the low divergence start in DminEmax strategy. Since DminEmax does not bias its first test to identify which PDF is most probable, the probability therefore, of the profile being healthy or tumor was calculated to be near 0.5 after the first test. As a dynamic strategy, however, it quickly converged and caught up on classification for the following tests.

From **Fig. 6d**, we successfully observed the binning and identification of a mock healthy profile using the 2-dimensional model. The CDAs were able to converge on the binning area over approximately four orders of magnitude. However, we do observe that on the fourth test that the two entropic-based dynamic strategies temporarily increased the binning area and had a decrease in information learned. This was due to the CDAs identifying and correcting an error made in one of the four previous tests resulting in consensus being split between two bins. As per the error correction method (**Supplementary Fig. 7**), the two disputed bins were merged to reestablish consensus causing an increase in the binning area and a drop in information learned. As these are both dynamic strategies, the next CDA was selected based on the merged bin and both strategies were able to recover information learned, correcting its previous error and continue to converge onto the biomarker.

After demonstrating successful convergence and classification using 2-dimensional PDF models, we extended these strategies to a 4-dimensional PDF model, based on the cross-multiplication of the PDFs for all four prognostic miRNAs. Again, for the mock tumor and healthy profiles (**Fig. 6e,f**) the dynamic strategies learned the most per test, converging the hypervolume the fastest, and correctly classifying the patient profiles in the fewest tests. The benefits of selecting CDAs optimized for high divergence are apparent with rapid classification, being able to identify a patient’s profile as tumor or healthy with a strong confidence in the first test. The capability of array parallelization and error correction, as demonstrated in **Fig. 6f**, enabled convergence of the hypervolume by over 3 orders of magnitude.

Ultimately dynamic strategies proved to be more effective in diagnosing patient profiles in fewer tests than the non-learning static methods. While Dmax is not necessarily the most efficient in converging on the binning area or hypervolume as other dynamic or static strategies because it is not optimizing for the most entropic CDAs, it is the most effective strategy to classify patient profiles. Due to Dmax and DmaxEmax testing a highly divergent CDA first, they classified patient profiles with considerable confidence in the first few tests, making these diagnostic strategies the most promising for PoC-aligned diagnosis. Dmax was also the fastest learning strategy achieving full convergence of information content 1-2 tests before the dynamic entropy methods and using significantly fewer tests than required by the static strategies.

## DISCUSSION

We have developed and characterized the CDA, a diagnostic device that recognizes concentration-dependent nucleic acid biomarkers. Analysis of CDA designs and experimental data revealed the key design parameters necessary for proper CDA function and yielded an effective op-amp comparator circuit model that related the thresholding properties of the devices to their domains. Using the characteristic thresholds of CDAs, we binned prognostic biomarkers for NSCLC. With each threshold defining the boundaries of a bin, CDA devices were demonstrated to consensus vote, error correct, and partition target biomarkers into defined concentration ranges (**Supplementary Fig. 7**). Parallel testing of samples with several CDAs allowed for the robust convergence towards smaller bins identifying biomarkers into concentration ranges with an AUC classification of 0.945-0.955 (**Fig. 5f**) at channel equilibrium and 0.892-0.954 in rapid 30-minute testing (**Fig. 5g**).

Motivated by the need to use minimal patient sample volume and reduce the number of tests required to achieve diagnosis geared for PoC-aligned platforms, we used a library of CDAs to demonstrate information theory-based diagnostic strategies. We concluded that the dynamic Dmax strategy, based on expectation maximization principles of utilizing highly divergent CDAs, was the most effective. Dmax was able to classify samples in a single test and learn the maximal amount of information content 1-2 tests faster than the other dynamic strategies.

Our success in harnessing information theory to improve the efficiency of molecular assays suggests that other core concepts from the field could prove useful for diagnostics. For instance, mutual information, which measures how much information learned about one variable can reduce uncertainty in another, could further advance diagnostic strategies^23^. Mutual information, as a metric, may be particularly useful for applications where we do not consider biomarkers as independent random variables and a test on one biomarker can reduce the number of tests required to resolve another. Information theory may also advance diagnostics through the development of purpose-built platforms that have a high channel capacity to relay a desired bits/unit time or bits/test rate. Channel capacity can provide a useful point of comparison between different systems to recognize relative diagnostic efficiency. Additionally, for complex diagnosis or large-scale omics screenings, the application of these core concepts from information theory may simplify and reduce unnecessary or non-informative testing and help identify and rank what biomarkers should be tested.

The classification of samples as healthy or cancerous was determined using a custom cohort from TCGA database (**Supplementary Table 3**). Collecting the open access miRNA biospecimen dataset for NSCLC and healthy control samples, we created PDF models to describe the probability of being in a certain health state as a function of miRNA concentrations. The custom cohort was limited however, as there are relatively low number of samples from healthy patients to build a high resolution PDF_Healthy_ model, most noticeable by the difference in Gaussian fit models R^2^ ranging from 0.865-0.949 for PDF_Healthy_ models and R^2^ = 0.986-0.990 for PDF_Tumor_ (**Supplementary Fig. 10**). Additionally, the cohort suffers from biases in the cancer stage, with over half of the NSCLC samples coming from stage I cancer patients, as well as limitations in patient demographic sampling.

Using high quality multidimensional PDFs to describe several health states of interest can enable the design of custom devices that target the most informative biomarkers. The advancement of an in-silico design code for CDAs, through the op-amp comparator circuit model can help guide the design of optimal entropic and divergent devices, as desired thresholds can be numerically determined based on the PDF models. We expect improvements in the op-amp circuit model will enable us to accurately design CDAs to activate at predefined and customized thresholds providing even more effective devices for the Dmax strategy. This approach to custom designing devices based on pre-existing PDF models can be generalized and applied to virtually any disease or health state of interest provided that the platform’s test results in a change of the expected probabilities of a target biomarker. This can be applied in multiple hypothesis testing, where we wish to create devices that best distinguish and classify multiple stages of cancer, or be able to rule-in rule-out various diseases that rely on common biomarkers. A major limitation towards advancing information theory-based devices is having detailed datasets to build PDF models from.

Use of CDAs and information theory-based Dmax strategy for PoC diagnosis of lung cancer will ultimately require finer resolution of the probability models with more dispersed demographic samplings. An increase in samples from other stages of lung cancer would also be beneficial to create a general classification model and PDFs for each cancer stage to track disease progression and effectiveness of treatments. Another area to be addressed with the PDF models is the effect of amplification techniques have on clinically derived miRNA samples. Prior work has demonstrated various linear and exponential amplification methods that are compatible with miRNAs. The gold-standard method for miRNA amplification is based on reverse transcription quantitative polymerase chain reaction using either stem-loop primers or a poly-A tail to improve compatibility with miRNAs^41^. Zhang et al. used an amplification method termed linear after the exponential PCR which was validated with the prognostic miRNAs −30a, −30d, - 148a, and −182^29^. Other demonstrated methods for miRNA amplification include isothermal techniques such as reverse transcription loop-mediated amplification, rolling circle amplification^43,44^, exponential amplification reaction^45^, and hairpin-mediated quadratic enzyme reaction^46^, amplifying miRNAs from the atto- and femto-molar concentrations into the nM - µM necessary for CDAs^41,47^.

Future developments of diagnostic devices and strategies can allow for not only generalizable detection of exogenous and endogenous nucleic acid biomarkers, but also for tracking disease progression and changes in biomarkers. Efficient strategies that minimize the number of tests needed to diagnose, support a future of PoC-aligned platforms where cancer and other highly intensive testing can be done outside of the clinic or centralized laboratories while using less invasive patient samples to achieve diagnosis. Beyond molecular diagnostics, an information theory-approach in nucleic acid nanotechnology can enhance and guide the development of DNA information storage and memory devices in designing optimized symbol codes, improve channel capacity, and error correction methods^22^. This can simplify designs by reducing the number of strands or tile components needed to transmit and relay information and address issues with noisy channels thereby improving confidence in a platform.

## MATERIALS AND METHODS

### CDA Fabrication

CDAs were designed using NUPACK software package (version 4.0.0.28) and DNA templates were purchased from IDT (Integrated DNA Technologies, Inc). Templates over 200 nucleotides in length were purchased in multiple DNA ultramer fragments and underwent PCR assembly using forward and reverse primers and ultramers (IDT) with Q5® High-Fidelity 2x Master Mix (New England Biolabs Inc., M0492L). Assembled templates were confirmed on a 1.5% agarose gel. If multiple bands appeared on the gel, then the correct length templates were gel purified (Promega, A9281). CDA templates were then transcribed using HiScribe® T7 High Yield RNA Synthesis kit (New England Biolabs Inc., E2040L) supplemented with 0.5 µL RNAse Inhibitor (New England Biolabs Inc., M0314L) per 20 µL reaction with 2 µL of template for at least 12 hours at 37 °C. Transcript products were purified with RNA cleanup purification kit (New England Biolabs Inc., T2040L). Concentration of purified RNA products were measured using NanoDrop UV-Vis Spectrophotometer (ThermoFisher Scientific, ND-ONEC-W) and diluted to a concentration of 10 µM in ultrapure water (Invitrogen^TM^, 10977015).

### Buffered Fluorophores Preparation

Buffered fluorophores were prepared and used for each experiment that had their respective cognate aptamer present: DFHBI-1T for Broccoli aptamer, DFHO for Corn aptamer, and YO3-3PEG-Biotin for Mango-IV aptamer.

For a 1 mL preparation of a 40 µM buffered DFHBI-1T, the following mixture was prepared on ice: 400 µL of 1 M HEPES buffer (Gibco^TM^, 15630080), 500 µL of 2 M KCl (Invitrogen^TM^, AM9640G), 50 µL of 1 M MgCl_2_ (Invitrogen^TM^, AM9530G), 2 µL of 20 mM DFHBI-1T (Lucerna, 410), and supplement with ultrapure water (Invitrogen^TM^, 10977015).

For a 1 mL preparation of a 20 µM buffered DFHO, the following reaction was prepared on ice: 400 µL of 1 M HEPES buffer (Gibco^TM^, 15630080), 500 µL of 2 M KCl (Invitrogen^TM^, AM9640G), 50 µL of 1 M MgCl_2_ (Invitrogen^TM^, AM9530G), 2 µL of 10 mM DFHO (Lucerna, 500), and supplement with ultrapure water (Invitrogen^TM^, 10977015).

For a 1 mL preparation of a 25 µM buffered YO3-3PEG-Biotin, the following reaction was prepared on ice: 400 µL of 1 M HEPES buffer (Gibco^TM^, 15630080), 500 µL of 2 M KCl (Invitrogen^TM^, AM9640G), 50 µL of 1 M MgCl_2_ (Invitrogen^TM^, AM9530G), 44 µL of 570 µM YO3-3PEG-Biotin (Applied Biological Materials Inc., G7957), and supplement with ultrapure water (Invitrogen^TM^, 10977015).

### CDA Reactions

All experiments were prepared on a 384-well plate (Corning, 3540) and read in a BioTek Synergy Neo2 multimode microplate reader (Agilent, BTNEO2) at 37 °C for 6 hours. The plate was linearly shaken for 30 seconds to ensure mixing before kinetic reading. 0.5 µM of purified CDA devices were tested with variable concentration of DNA oligo target biomarkers (IDT) and buffered fluorophores. For CDAs that had Broccoli, Corn, or Mango-IV aptamers present, their respective buffered fluorophores were added at final concentration of 2 µM of DFHBI-1T, 1 µM of DFHO, and 2 µM of YO3-3PEG-Biotin unless otherwise noted. Reactions were set to 35 µL and supplemented with ultrapure water (Invitrogen^TM^, 10977015).

Broccoli/DFHBI-1T channel was read at an excitation/emission wavelength 472 nm and 507 nm. Corn/DFHO channel was read at an excitation/emission wavelength 505 nm and 545 nm. Mango-IV/YO3-3PEG-Biotin channel was read at an excitation/emission wavelength of 595 nm and 620 nm. All wavelengths were read with a bandwidth of ±10 nm.

For all experiments unless otherwise specified, a no-target control containing the CDA and respective fluorophores was used along with a background control containing only the fluorophores. For the concentration series experiment the biomarker concentrations were tested at 0.01, 0.02, 0.03, 0.04, 0.05, 0.1, 0.2, 0.3, 0.4, 0.5, 1, 2, 3, 4, 5, 10, 20, 30, 40, and 50 µM. All experimental groups were repeated in technical triplicates. Analysis of plate reader experiments were done at the 6-hour endpoint, unless otherwise noted, to ensure all channels had reached equilibrium.

### Fluorescent Imaging

CDA reactions were incubated at 37°C for 10 minutes and then illuminated on a blue light source and orange filter with Safe Imager^TM^ 2.0 Blue-Light Transilluminator (ThermoFisher, G6600). Images were taken from an iPhone 17 Pro camera. The concentration of DFHBI-1T and DFHO were set to 2 µM and 3 µM respectively. The concentration of biomarkers to activate fluorescence in bins 1, 2, and 3, were set to 0.25 µM, 0.75 µM, and 10 µM respectively.

### TCGA Custom Cohort for studying NSCLC

A custom cohort to study NSCLC cases was created from open access TCGA biospecimen datasets. In total there were 739 tumor, and 64 healthy aliquots recorded in the cohort. See **Supplementary Table 3**, for cohort details. The PDFs developed from the custom cohort were least-squares fitted to Gaussian distributions. Construction of 2-dimensional (**Supplementary Fig. 13**) and 4-dimensional joint PDF models used in **Fig. 6** were calculated by cross multiplying 1-dimensional PDF models from **Supplementary Fig. 12**. Joint PDF models assume that the concentrations of each miRNA are independent random variables.

### Statistical Methods and Additional Calculations

Plate reader fluorescence and figures were analyzed and prepared using Python Jupyter Notebook (version 6.5.4), Microsoft Excel, and Adobe Illustrator. Averages from technical triplicates were calculated as the arithmetic mean. Error bars or shaded regions on plots unless otherwise indicated represent ±1 standard deviation from the mean. Fold change was measured as the fluorescence signal of the experimental group divided by the fluorescence signal of the no-target control. R^2^ indicates the coefficient of determination, r indicates Pearson correlation, and ρ indicates Spearman rank correlation. Welch’s T-Test was used to determine statistical significance and report p-values between histograms of miRNA read counts from TCGA custom cohort.

The length of bins, measured in µM, was calculated as the difference from the identified upper and lower bound thresholds, where the maximum upper and minimum lower bound thresholds were 50 µM and 0 µM respectively. The area of a bin in a 2-dimensional joint PDF, measured in µM^2^, was calculated by the multiplication of the two bin lengths each defined by the independent binning of two miRNAs. The hypervolume of a bin in a 4-dimensional joint PDF, measured in µM^4^, was calculated by the multiplication of the four bin lengths from the independent binning of all four prognostic miRNAs.

### Data Availability

See **Supplementary Table 4**, for all CDA and target biomarker sequences. Experimental data are available from the corresponding author on reasonable request.

## Data Availability

See Supplementary Table 4, for all CDA and target biomarker sequences. Experimental data are available from the corresponding author on reasonable request.

## Acknowledgments

This work was supported by startup funds from Boston University; Defense Advanced Research Projects Agency (DARPA) funding (Contract No. N66001-23-2-4042); and a National Institutes of Health (NIH) U01 award (1U01AI148319-01), and R01 award (1R01EB031893) to A.A.G. The views, opinions and/or findings expressed are those of the authors and should not be interpreted as representing the official views or policies of the Department of Defense or the U.S. Government. The content is solely the responsibility of the authors and does not necessarily represent the official views of the National Institutes of Health.

## Author contributions

A.E. performed wet lab experiments, design of CDAs, data analysis, and simulations. A.E. and A.A.G. developed CDA designs. A.E. and A.A.G. interpreted data and wrote manuscript.

## Declaration of interests

A.A.G. is a cofounder of En Carta Diagnostics Inc. and Gardn Biosciences. The authors declare no other competing interests.

## Notes

### Author Declarations

This study used ONLY openly available human data that were originally located at The Cancer Genome Atlas (TCGA) database.

