## Supplementary Information for "Information Theory-Guided Detection of Biomarkers Using Programmable Aptamer Arrays"

**Supplementary Note 1: Operational Amplifier Comparator Circuit Modeling**

The operational amplifier (op-amp) comparator circuit model, as illustrated in **Figure 4** uses two distinctive circuit components: the op-amp and voltage divider. The op-amp solves the following inequality and provides a conditional output:

$$\begin{cases} If\ V_{IN} > V_{REF,CH}\ then\ V_{OUT,CH} = +V_{CC} \\ If\ V_{IN} < V_{REF,CH}\ then\ V_{OUT,CH} = -V_{CC} \end{cases}$$

Where $V_{REF,CH}$ and $V_{IN}$ are the input voltages at the negative and positive nodes respectively, and $+V_{CC}$ and $-V_{CC}$ are the op-amp output rail voltages.

To model a 2-channel CDA, there are two op-amps and consequently, a $V_{REF,CH1}$ and $V_{REF,CH2}$ present at the negative input nodes for each op-amp to describe $V_{OUT,CH1}$ and $V_{OUT,CH2}$ respectively. $V_{REF,CH1}$ and $V_{REF,CH2}$ in-turn are governed by the voltage divider equations:

$$V_{REF,CH1} = V_{REF} \times \left(\frac{R_1}{R_1 + R_2 + R_3}\right)$$

$$V_{REF,CH2} = V_{REF} \times \left(\frac{R_1 + R_2}{R_1 + R_2 + R_3}\right)$$

Over a voltage sweep of $V_{IN}$, represented as the voltage across the $R_{LOAD}$ resistor is akin to the target biomarker concentration, while $V_{REF,CH1}$ and $V_{REF,CH2}$ are recognized to be analogous to $Threshold_{CH1}$ and $Threshold_{CH2}$. $V_{REF}$ can be conceptualized as an innate maximum threshold, as according to the voltage divider equations, $V_{REF,CH1}$ and $V_{REF,CH2}$ do not exceed $V_{REF}$. $V_{OUT,CH1}$ and $V_{OUT,CH2}$ become analogous to the output channel fluorescence for low concentration CH1 and high concentration CH2 respectively. $+V_{CC}$ becomes the maximum normalized fluorescence observed when a channel is saturated with biomarkers and $-V_{CC}$ is the normalized fluorescence observed by that channel when no biomarkers are present.

To further connect the circuit analogy to the design features of the CDA, we set the resistors $R_1$, $R_2$, and $R_3$ to be functions of the domain lengths **TH1**, **TH2**, and **B** and **C**, respectively. **TH1** represents the binding site necessary to initiate the activation of CH1, as such, the “resistance” needed to define the numerator of the $V_{REF,CH1}$ equation, $R_1$, will be linked to **TH1**. Similarly, for $R_2$, CH2 only activates with base-pair binding of the biomarker to **TH2** which happens after CH1 has been activated overcoming $R_1$ resistance. $R_3$ was set to be a function of the **B** and **C** domains because both CH1 and CH2 have the same **B** and **C** domain lengths in their hairpins and consequently their influence through the toehold-mediated strand-displacement process for each channel would similarly affect $Threshold_{CH1}$ and $Threshold_{CH2}$.

To elucidate an exact relationship between the resistors and domain, the domain lengths of 31 functional 2-channel CDA devices were modeled to simultaneously solve the two voltage divider equations. Various combinations of parameterizing the resistor equations as a function of the domain lengths were tested. Resistor equations were evaluated using gradient descent and the formulation that best minimized the residuals sum squared (RSS) between the voltage model and the observed thresholds was selected.

$$RSS = \sum \left(Threshold_{CH1} - V_{REF,CH1}\right)^2 + (Threshold_{CH2} - V_{REF,CH2})^2$$

Additional consideration was given to balance how many fitting parameters were used for the resistor equations to avoid unnecessary terms and overfitting the data. As such, with 31 CDAs providing 124 domain lengths (31x4) to fit 62 threshold values (31x2), we desired to not use more than 3 fitting parameters in our entire model. The final equations for the resistors were determined to be:

$$R_1(TH1) = \frac{1}{length(TH1) + a}$$

$$R_2(TH2) = \frac{1}{length(TH2)}$$

$$R_3(B, C) = \frac{k}{length(B)} + length(C)$$

Gradient descent was repeated numerous times with random starting values of the fitting parameters to ensure that the RSS minimum converged to consistent parameter values. A mapping of the RSS as a function of the “a” and “k” parameters confirmed a single minimum (see plot below). All modeled formulations of the resistor equations that allowed any $V_{OUT,CH1}$ or $V_{OUT,CH2}$ to have a negative value were discounted, and shown in the RSS mapping with a value >25.

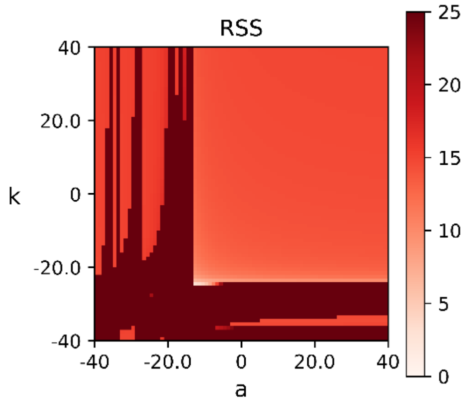

**Residual Sum Square Mapping of Circuit Model**

Besides the “a” and “k” from the resistor equations, an additional $V_{NOISE}$ fitting parameter was included, modifying the final fitted voltage divider equations.

$$V_{REF,CH1} = V_{REF} \times \left( \frac{R_1(TH1)}{R_1(TH1) + R_2(TH2) + R_3(B,C)} \right) + V_{NOISE}$$

$$V_{REF,CH2} = V_{REF} \times \left( \frac{R_1(TH1) + R_2(TH2)}{R_1(TH1) + R_2(TH2) + R_3(B,C)} \right) + V_{NOISE}$$

With the inclusion of “a”, “k”, and $V_{NOISE}$ parameters, $V_{REF}$ was not fitted and kept to a default value of 1 volt. See, **Supplementary Table 2** for all parameter values.

**Supplementary Note 2: Information Theory Approach to Concentration Detector Arrays**

Three core concepts from information theory were applied to CDAs for the development of efficient diagnostic strategies: information content, Shannon entropy (H), and Kullback-Leibler divergence (D)[1,2].

Information content quantitatively represents the amount of information learned about a biomarker once it is identified into a particular bin by a CDA test. It is based on the associated probability of a certain health state defined in the range of that bin. A bin, measured in $\mu$M, is defined by its upper and lower bound thresholding concentrations. The probability of a biomarker represented by the variable "y", being in a certain health state within a bin "x" is calculated by the definite integral of the associated health state PDF (healthy or tumor).

$$p(x) = \int_{Lower\ Bound\ Threshold}^{Upper\ Bound\ Threshold} PDF(y)\ dy$$

Information content, measured in bits for a bin, is defined per health state as:

$$Information\ Content(x) = -log_2(p(x))$$

1 bit represents bisecting a PDF in half. Intuitively, information can be considered as a metric of uncertainty uncovered. A completely deterministic state or event would require 0 bits of information to be learned as there is no uncertainty associated with that state of event happening[3]. A highly probable event would have a low information content, as we would not be surprised to see that state or event occurring. As the probability of a certain state or event decreases, such as a biomarker being in a certain concentration range, the amount of information learned increases. In analysis, we assume that each miRNA acts as an independent random variable and consequently, the information learned by binning each miRNA is additive.

Shannon entropy represents the expected amount of information that is to be learned from a particular CDA test. A low entropic device represents a binning by the CDA that heavily biases the probability of a health state towards a particular bin, and as such, there is a low surprisal when identifying a biomarker in that biased bin. Entropy, measured in bits, over discretized probability distribution for a set of X bins, is defined as:

$$H(X) = -\sum_{x \in X} p(x) \times log_2(p(x))$$

Entropy is maximized under a uniform distribution of uncertainty[1,3,4]. Given that a 2-channel CDA creates three bins, the maximum entropy a 2-channel device can achieve is when each bin discretizes a PDF into three 1/3 probabilities, attaining a maximum entropy of 1.58 bits.

$$X = [x_1, x_2, x_3]$$

$$p(X) = [\frac{1}{3}, \frac{1}{3}, \frac{1}{3}]$$

$$H(X) = -\left[\frac{1}{3} \times log_2\left(\frac{1}{3}\right) + \frac{1}{3} \times log_2\left(\frac{1}{3}\right) + \frac{1}{3} \times log_2\left(\frac{1}{3}\right)\right]$$

$$H(X) = 1.58\ bits$$

As observed in **Supplementary Figure 14**, for a library of CDAs, some do achieve near maximal entropy for both PDF models. With the most entropic device tested having a Shannon entropy of 1.55 bits.

Kullback-Leibler divergence represents the amount of excess information to be learned if we incorrectly assume the wrong PDF model. D serves as a pseudo-distance calculation between the binning of $PDF_{Healthy}$ and $PDF_{Tumor}$ for a given set of bins X.

$$D(p_{Healthy} || p_{Tumor}) = \sum_{x \in X} p_{Healthy}(x) \times log_2\left(\frac{p_{Healthy}(x)}{p_{Tumor}(x)}\right)$$

A high divergence may bias and maximize $PDF_{Healthy}$ into one bin and $PDF_{Tumor}$ into another. Conversely, a low divergence represents a discretization by the CDA that does not bias one PDF over another, and from the perspective of the CDA, the discretized PDFs are very similar. A divergence of 0 bits represents a device that discretizes both PDFs in such a way that the probability of being healthy or tumor per bin is identical.

All calculations of information content, H, and D are based on the PDF models constructed in **Supplementary Figure 12**.

### Supplementary Figures

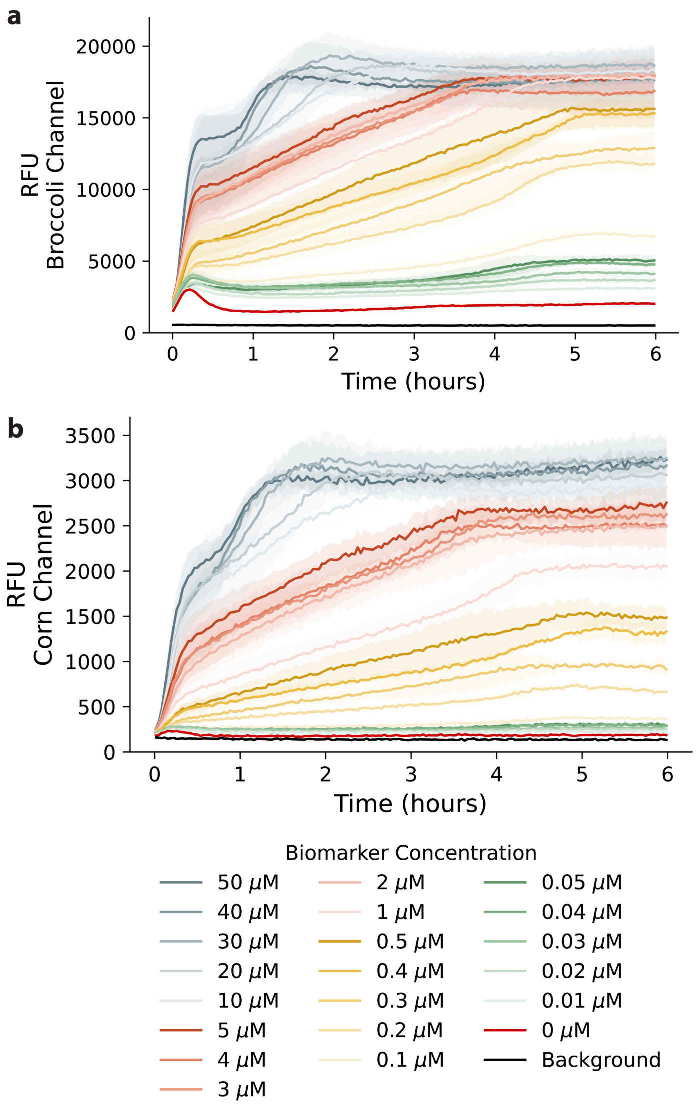

**Supplementary Fig. 1 | Time kinetics for a representative 2-channel CDA.**

**a**, Fluorescence (RFU) measurement of Broccoli channel, CH1 across biomarker concentration series. Excitation/emission wavelengths set to 472, 507 nm.

**b**, Fluorescence (RFU) measurement of Corn channel, CH2 across biomarker concentration series. Excitation/emission wavelengths set to 505, 545 nm.

N = 3. Solid line indicates mean, shaded region indicates ±1 standard deviation from the mean.

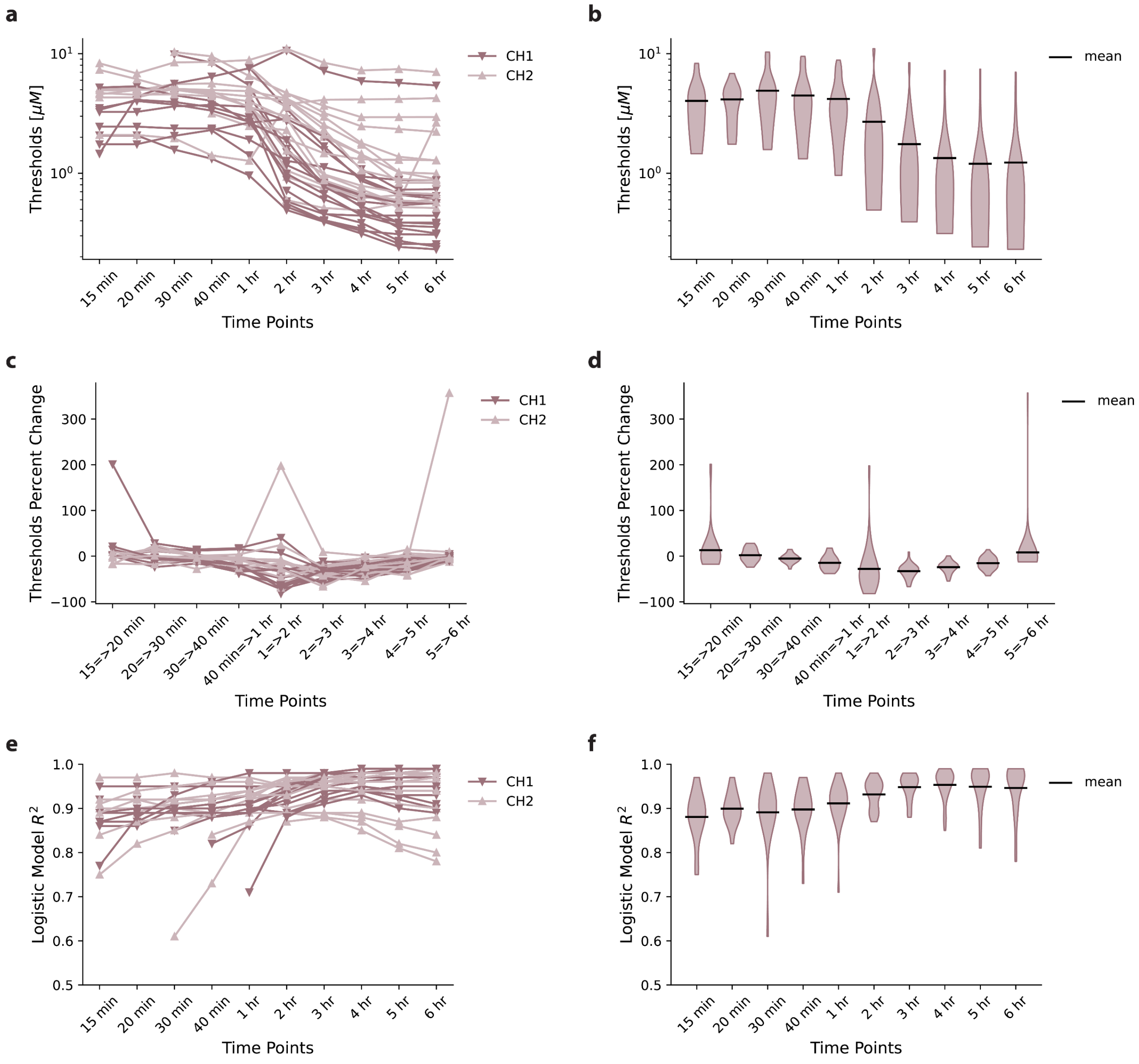

**Supplementary Fig. 2 I Time-dependent threshold analysis of miRNA targeting CDAs.**

**a**, Channel thresholds trace of individual CDAs over time beginning at the earliest timepoint a threshold can be calculated for each channel.

**b**, Violin plot of thresholds over time.

**c**, Threshold percent change trace of individual CDAs over time.

**d**, Violin plot of threshold percent change over time.

**e**, Logistic model $R^2$ values used to identify thresholds from individual CDAs over time.

**f**, Violin plot of logistic model $R^2$ values used to identify thresholds.

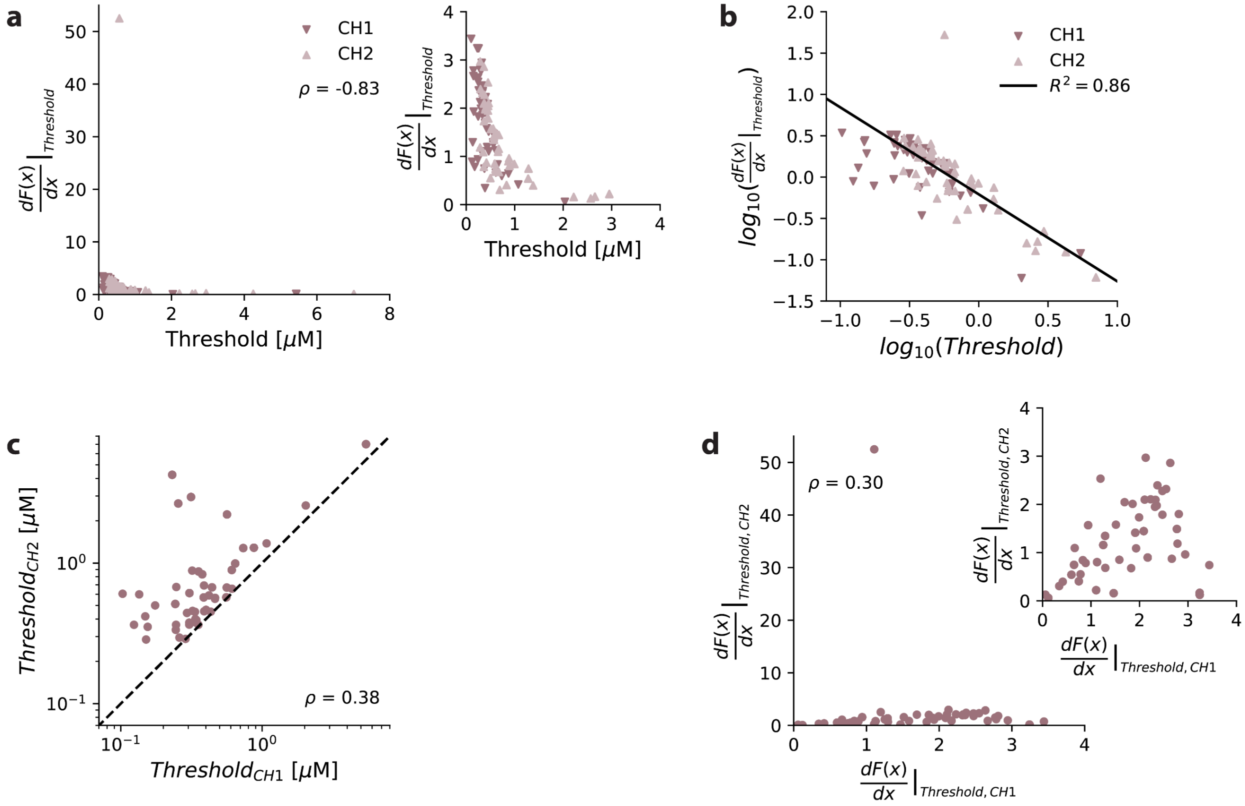

**Supplementary Fig. 3 | Threshold-derivative relationship for all CDAs.**

**a**, Logistic model derivative at the channel's thresholding concentration as a function of the channel's thresholding concentration. Spearman rank correlation ρ = -0.83.

**b**, Logarithmic relationship observed in part **a** with linear regression fit (solid line). Coefficient of determination $R^2 = 0.86$ for regression fit.

**c**, Thresholding concentration of CH1 versus CH2. Functional CDAs are above dashed line. Spearman rank correlation ρ = 0.38.

**d**, Derivative of logistic model for CH2 at $Threshold_{CH2}$ versus derivative of logistic model for CH1 at $Threshold_{CH1}$. Spearman rank correlation ρ = 0.30.

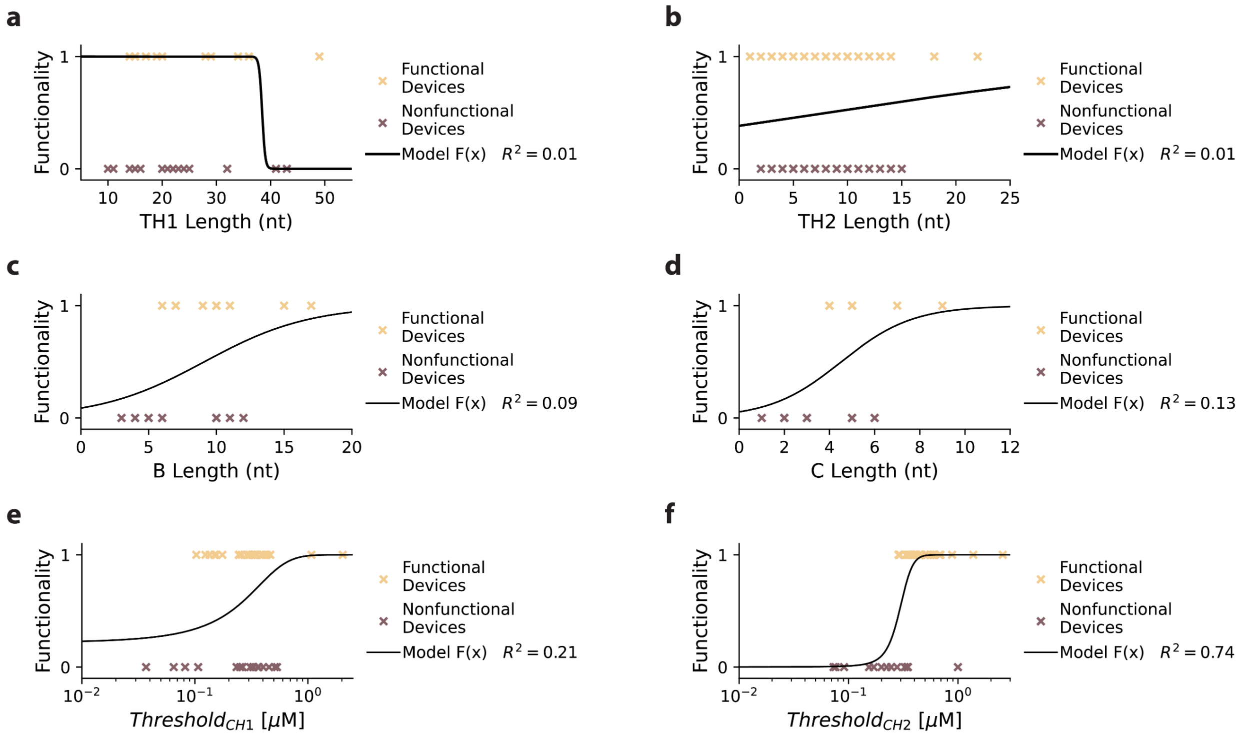

**Supplementary Fig. 4 I CDA functionality across key design features.**

**a**, Length of **TH1** domain in nucleotides versus device labeled functionality. Logistic regression model fitted with $R^2 = 0.01$. Sensitivity, specificity, and accuracy for classifying functionality 0.97, 0.07, and 0.52, respectively.

**b**, Length of **TH2** domain in nucleotides versus device labeled functionality. Logistic regression model fitted with $R^2 = 0.01$. Sensitivity, specificity, and accuracy for classifying functionality 0.52, 0.61, and 0.57, respectively.

**c**, Length of **B** domain in nucleotides versus device labeled functionality. Logistic regression model fitted with $R^2 = 0.09$. Sensitivity, specificity, and accuracy for classifying functionality 0.81, 0.32, and 0.57, respectively.

**d**, Length of **C** domain in nucleotides versus device labeled functionality. Logistic regression model fitted with $R^2 = 0.13$. Sensitivity, specificity, and accuracy for classifying functionality 0.84, 0.36, and 0.60, respectively.

**e**, $Threshold_{CH1}$ in $\mu$M versus device labeled functionality. Logistic regression model fitted with $R^2 = 0.21$. Sensitivity, specificity, and accuracy for classifying functionality 0.77, 0.58, and 0.68, respectively.

**f**, $Threshold_{CH2}$ in $\mu$M versus device labeled functionality. Logistic regression model fitted with $R^2$ = 0.74. Sensitivity, specificity, and accuracy for classifying functionality 0.94, 0.87, and 0.90, respectively.

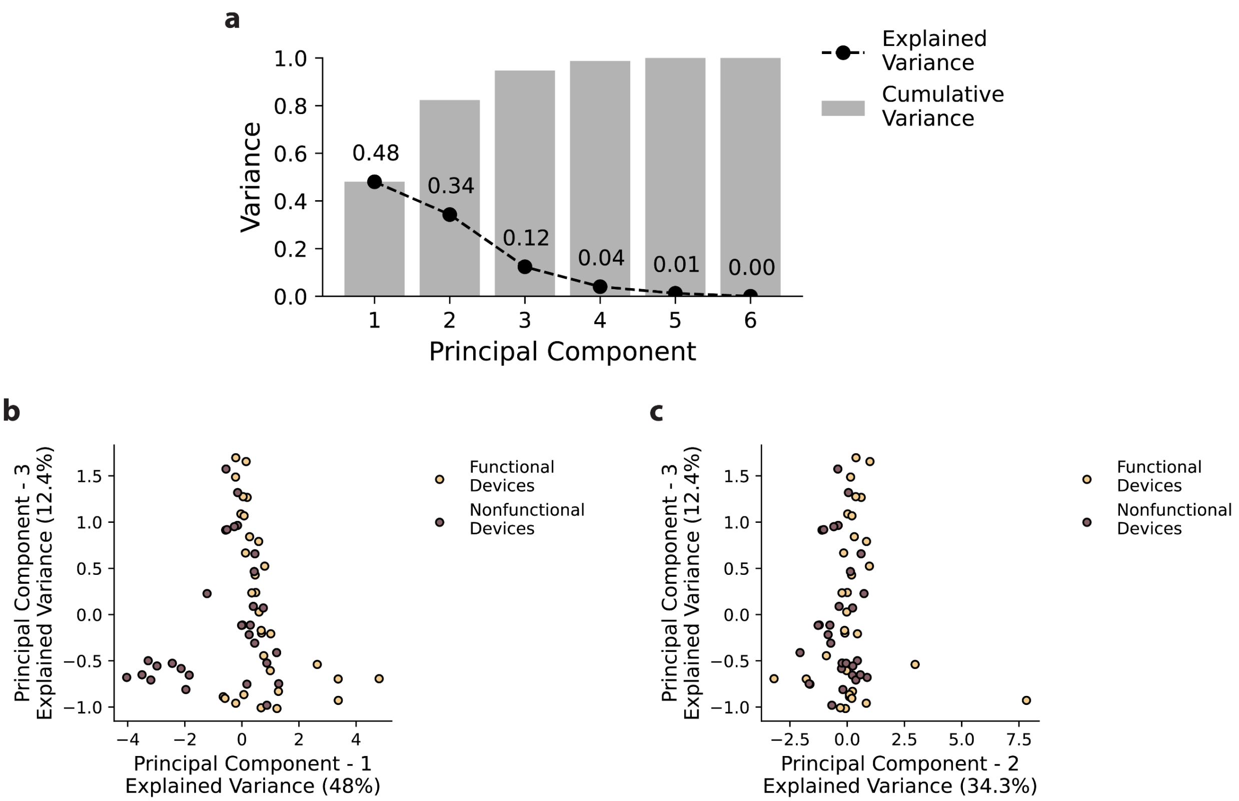

**Supplementary Fig. 5 I Principal component analysis for device functionality.**

**a**, Scree plot of explained variance per principal component.

**b**, Principal component 1 versus principal component 2 of device functionality.

**c**, Principal component 2 versus principal component 3 of device functionality.

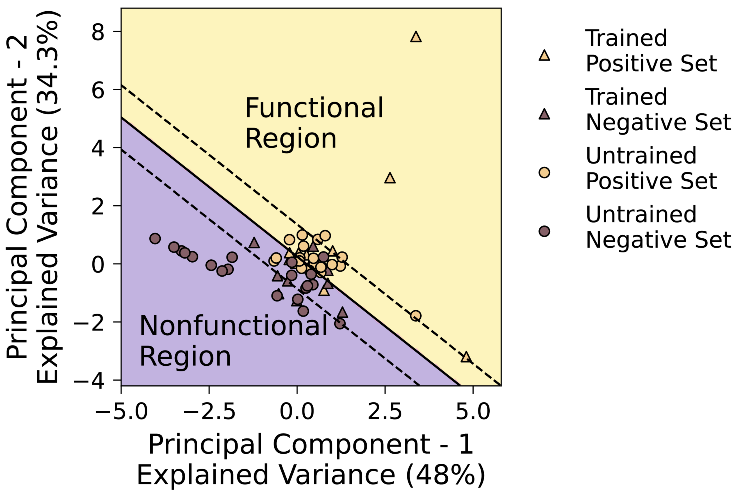

**Supplementary Fig. 6 | Robustness of decision boundary functionality classifier.**

Linear decision boundary classifier made with a training set of randomly selected 10 functional (positive) and 10 nonfunctional (negative) devices. Classification sensitivity, specificity, and accuracy of the remaining 42 (21 positive and 21 negative) devices are 90.5%, 85.7%, and 88.1% respectively. Solid line indicates the 0-contour line of the decision boundary curve, the upper and lower dashed lines indicate the 0.95 and -0.95 contour lines of the decision boundary curve between classifying functional devices labeled as value 1 and nonfunctional devices labeled with value of -1.

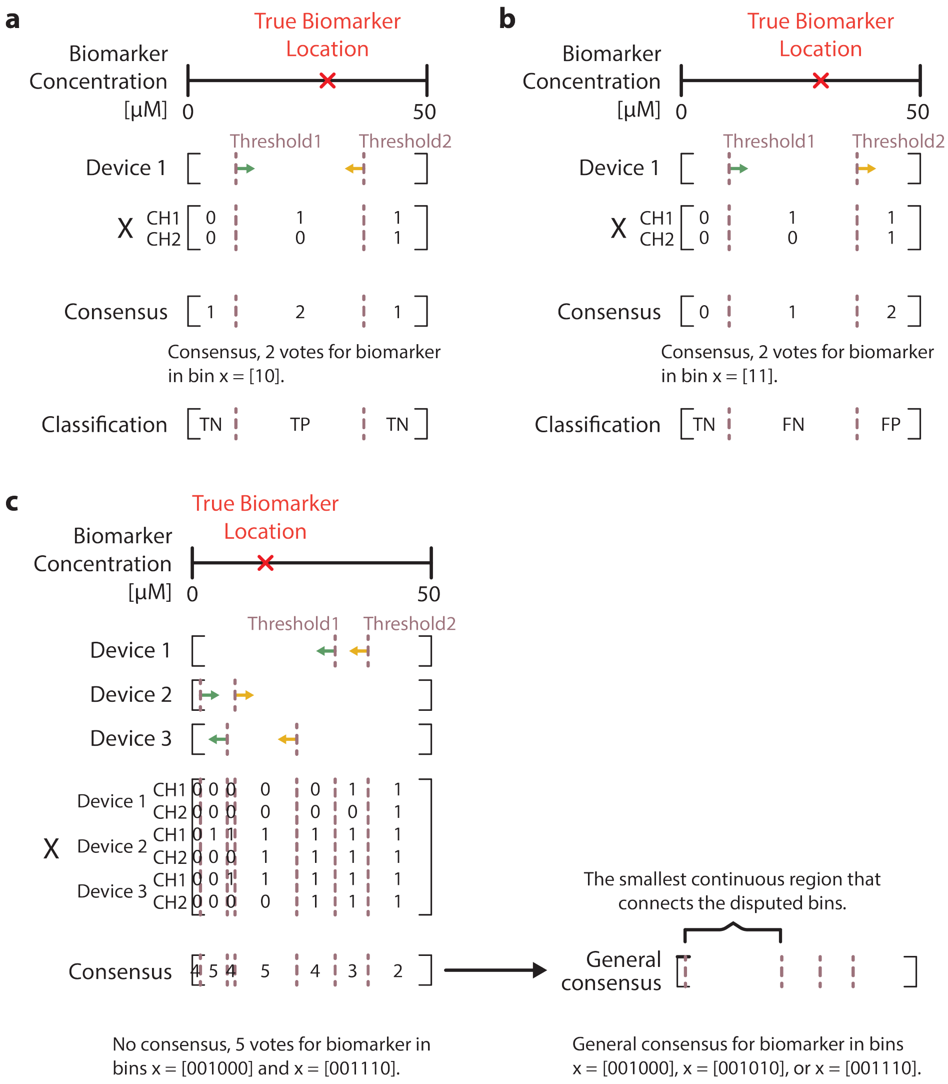

**Supplementary Fig. 7 | Consensus voting and error correction.**

**a**, Correct classification of biomarker into CDA bins through consensus voting.

**b**, Incorrect classification of biomarker into CDA bins through consensus voting.

**c**, Error correction method when consensus voting results in a dispute between two bins.

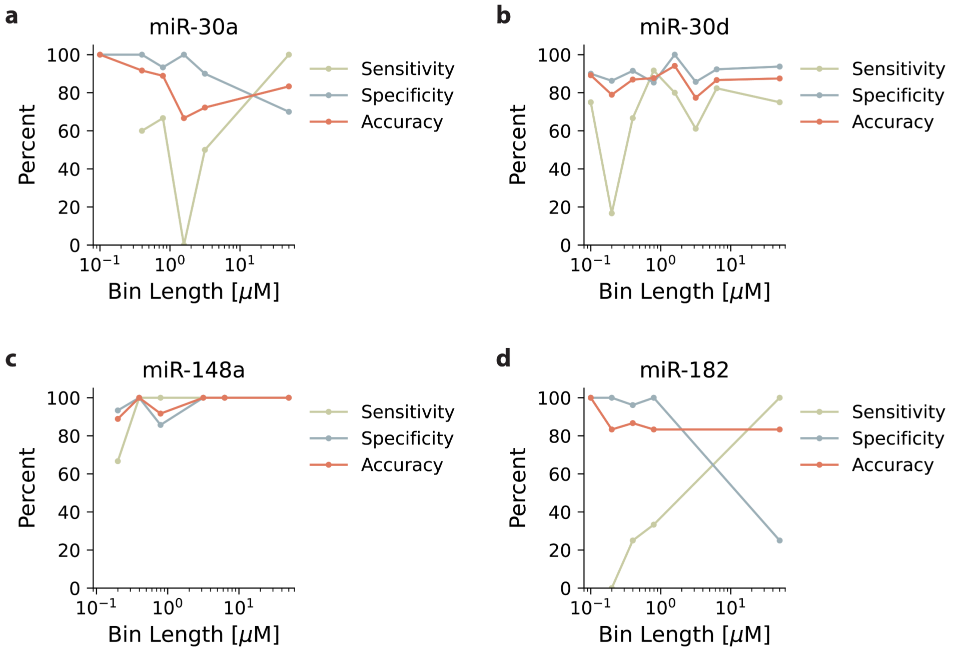

**Supplementary Fig. 8 | CDA binning per prognostic miRNA.**

**a**, Bin classification sensitivity, specificity, and accuracy of library of one and two parallel CDAs targeting miR-30a, as a function of bin length.

**b**, Bin classification sensitivity, specificity, and accuracy of library of one and two parallel CDAs targeting miR-30d, as a function of bin length.

**c**, Bin classification sensitivity, specificity, and accuracy of library of one and two parallel CDAs targeting miR-148a, as a function of bin length.

**d**, Bin classification sensitivity, specificity, and accuracy of library of one and two parallel CDAs targeting miR-182, as a function of bin length.

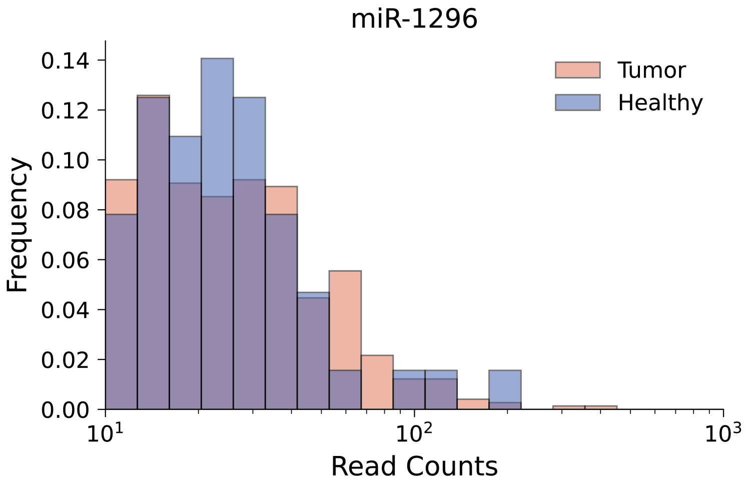

**Supplementary Fig. 9 l Histogram read count for miR-1296 from NSCLC and healthy patient sample aliquots.**

Data taken from TCGA custom cohort (see **Supplementary Table 3**) miRNA read counts. Histogram plotted on log-spaced bins. miR-1296 identified to have invariant expression profile for tumor and healthy sample status. $N_{Tumor} = 739$, $N_{Healthy} = 64$, Welch's T-test p-value = 0.92.

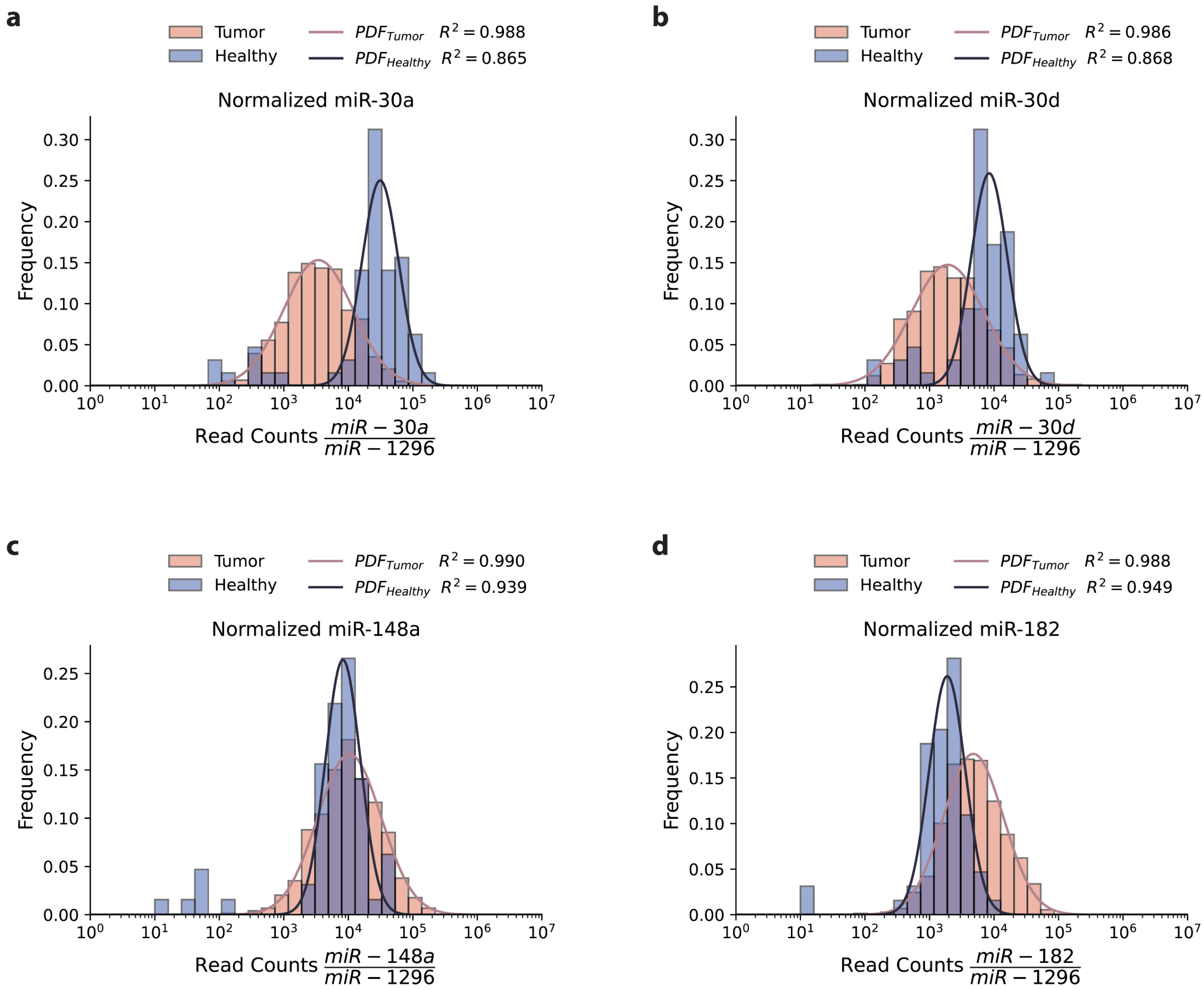

**Supplementary Fig. 10 I Histogram normalized read counts and probability distribution functions for prognostic miRNAs for NSCLC.**

**a**, Histogram and Gaussian least-squares fitted probability distribution functions for normalized miR-30a read counts. $R^2$ = 0.988 for fitted tumor PDF and $R^2$ = 0.865 for fitted healthy PDF. Welch's T-test p-value = $4.4 \times 10^{-10}$.

**b**, Histogram and Gaussian least-squares fitted probability distribution functions for normalized miR-30d read counts. $R^2$ = 0.986 for fitted tumor PDF and $R^2$ = 0.868 for fitted healthy PDF. Welch's T-test p-value = $5.8 \times 10^{-7}$.

**c**, Histogram and Gaussian least-squares fitted probability distribution functions for normalized miR-148a read counts. $R^2$ = 0.990 for fitted tumor PDF and $R^2$ = 0.939 for fitted healthy PDF. Welch's T-test p-value = $6.1 \times 10^{-9}$.

**d**, Histogram and Gaussian least-squares fitted probability distribution functions for normalized miR-182 read counts. $R^2 = 0.988$ for fitted tumor PDF and $R^2 = 0.949$ for fitted healthy PDF. Welch's T-test p-value = $5.2 \times 10^{-38}$.

Data taken from custom TCGA cohort (see **Supplementary Table 3**) miRNA read counts. $R^2$ indicates the coefficient of determination correlation. $N_{Tumor} = 739$ and $N_{Healthy} = 64$.

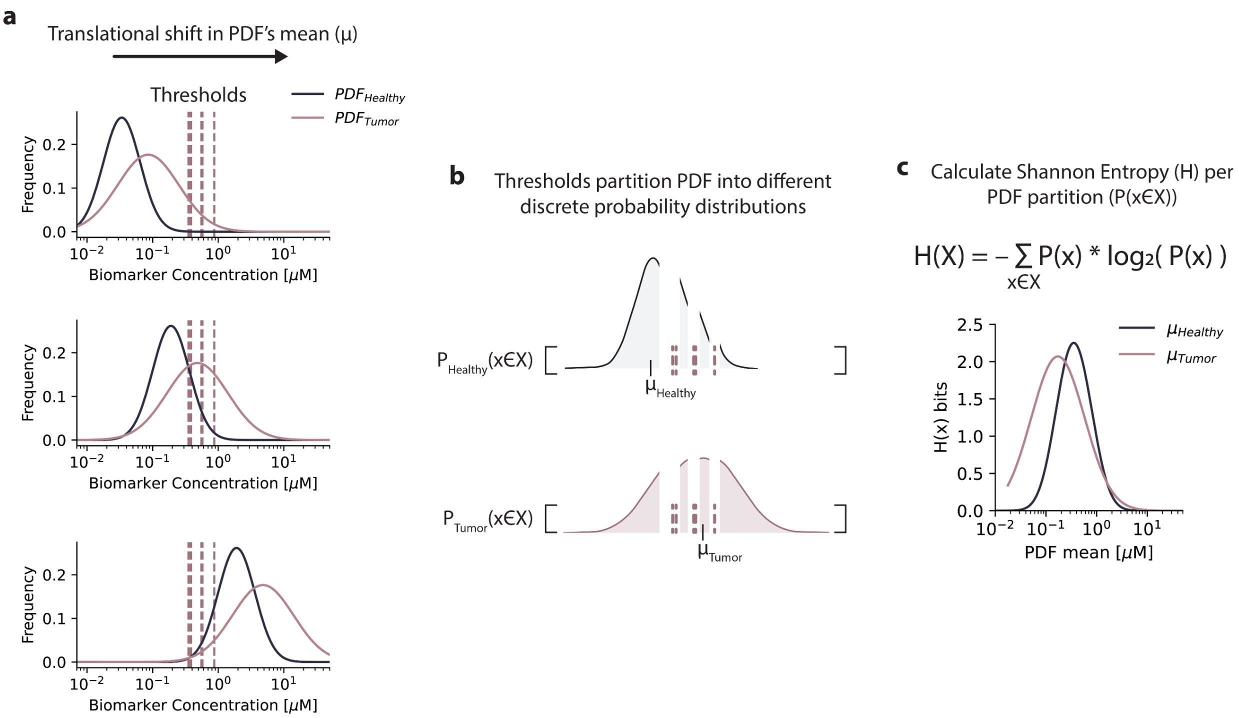

**Supplementary Fig. 11 | Identifying optimized amplification of PDF using Shannon entropy.**

**a**, A pair of healthy and tumor normalized PDFs for a prognostic miRNA, translationally shifted across biomarker concentration x-axis into the effective concentration range of a library of CDA thresholds. Assumes "amplification" does not bias relative concentration distribution.

**b**, For each position of the PDF distributions centered at different mean values, distributions are partitioned across the thresholding concentration values based on a library of CDAs. The probabilities for each PDF per bin created by the thresholds are calculated by the definite integral of the PDF defined by each bin's range.

**c**, The Shannon entropy is calculated for each discrete probability distribution created by the binning of the PDF into numerous bins. Shannon entropy given a library of CDA devices versus the mean value of the PDF functions allow for the identification of the most entropic concentration the PDFs should be amplified to.

Maroon dashed lines indicate the thresholding concentrations for a library of CDAs.

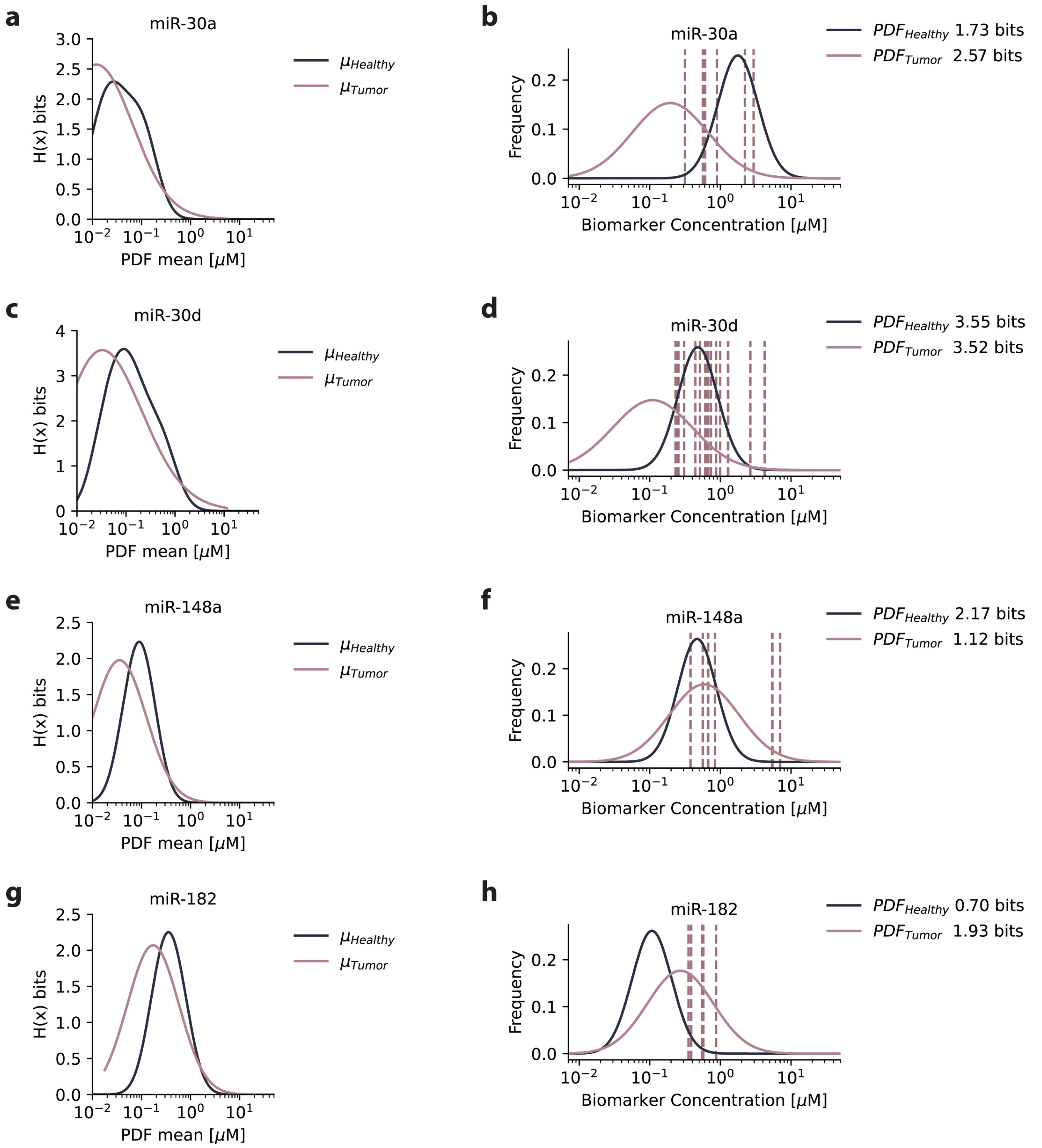

**Supplementary Fig. 12 I Normalized probability distribution functions for prognostic miRNAs at optimized entropic concentration for CDA binning.**

**a**, **c**, **e**, and **g**, Shannon entropy of healthy and tumor PDFs centered at mean values given a library of CDAs targeting each prognostic miRNAs.

**b**, **d**, **f**, and **h**, Healthy and tumor PDF for all 8 models were simultaneously optimized to the maximum Shannon entropy measured in bits, such that each model was translationally “amplified” by the same factor. Maroon dashed lines indicate the thresholding concentrations for library of CDAs targeting that specific miRNA.

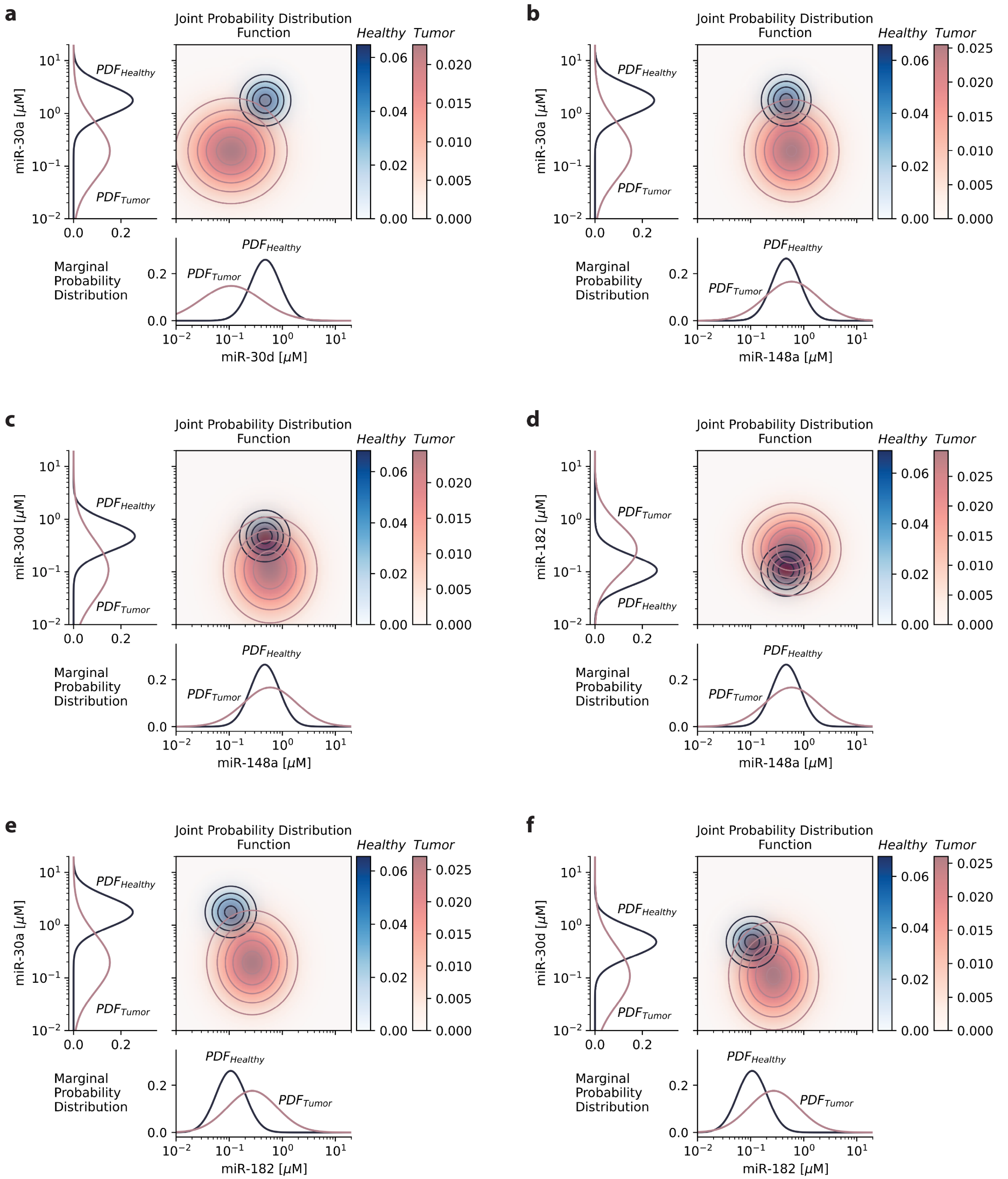

**Supplementary Fig. 13 | Amplified joint PDFs for normalized prognostic miRNAs for NSCLC.**

**a**, Joint normalized PDFs for miR-30d and miR-30a.

**b**, Joint normalized PDFs for miR-148a and miR-30a.

**c**, Joint normalized PDFs for miR-148a and miR-30d.

**d**, Joint normalized PDFs for miR-148a and miR-182.

**e**, Joint normalized PDFs for miR-182 and miR-30a.

**f**, Joint normalized PDFs for miR-182 and miR-30d.

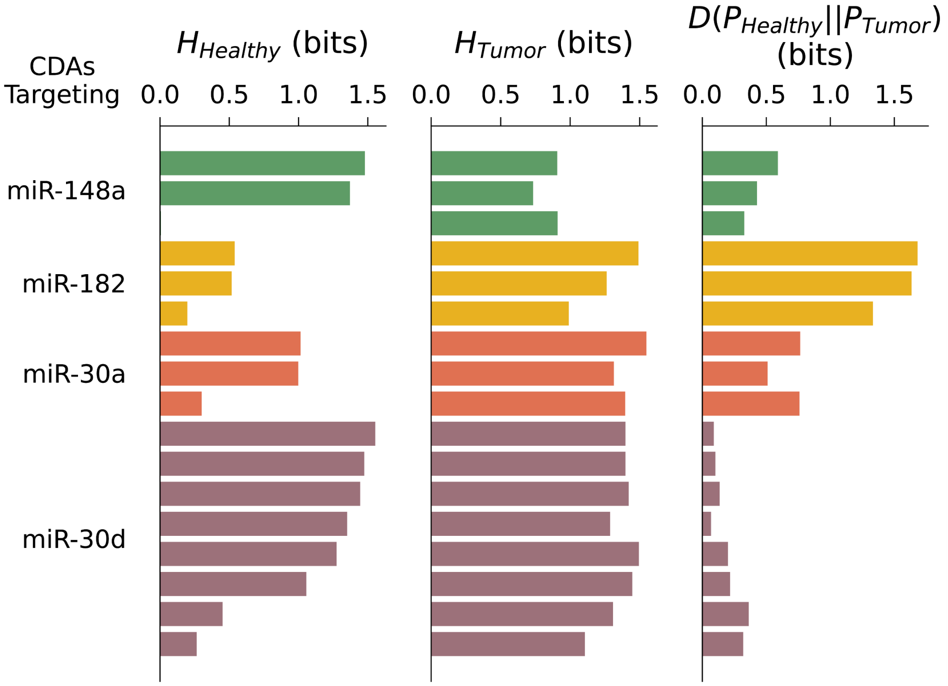

**Supplementary Fig. 14 | Shannon entropy and Kullback-Leibler divergence for library of miRNA targeting CDAs.**

The Shannon entropy assuming the amplified $PDF_{Healthy}$ model ($H_{Healthy}$) and $PDF_{Tumor}$ model ($H_{Tumor}$), and Kullback-Leibler divergence (D) calculated for a library of miRNA targeting 2-channel CDAs with characterized thresholds that discretize the PDFs identified in **Supplementary Figure 12**.

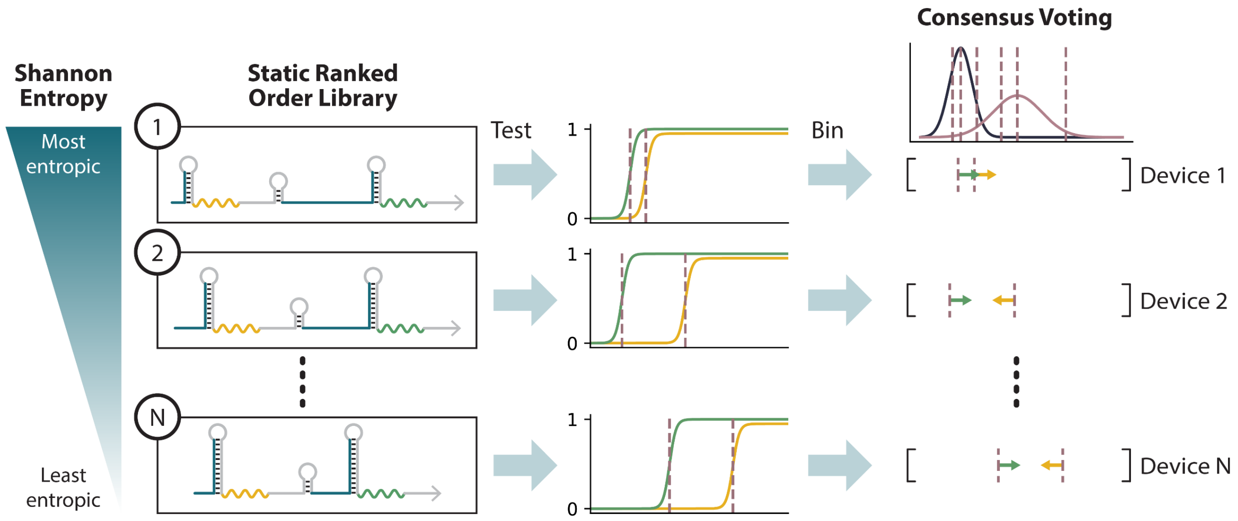

**Supplementary Fig. 15 I Static information theory-based diagnostic strategies.**

Static rank order diagnostic strategy. Rank library of CDAs according to pre-test Shannon entropy based on either $PDF_{Healthy}$ or $PDF_{Tumor}$ and test devices in descending entropy. Entropy of the remaining library is not recalculated or considered based on the binning outputs of previous tests.

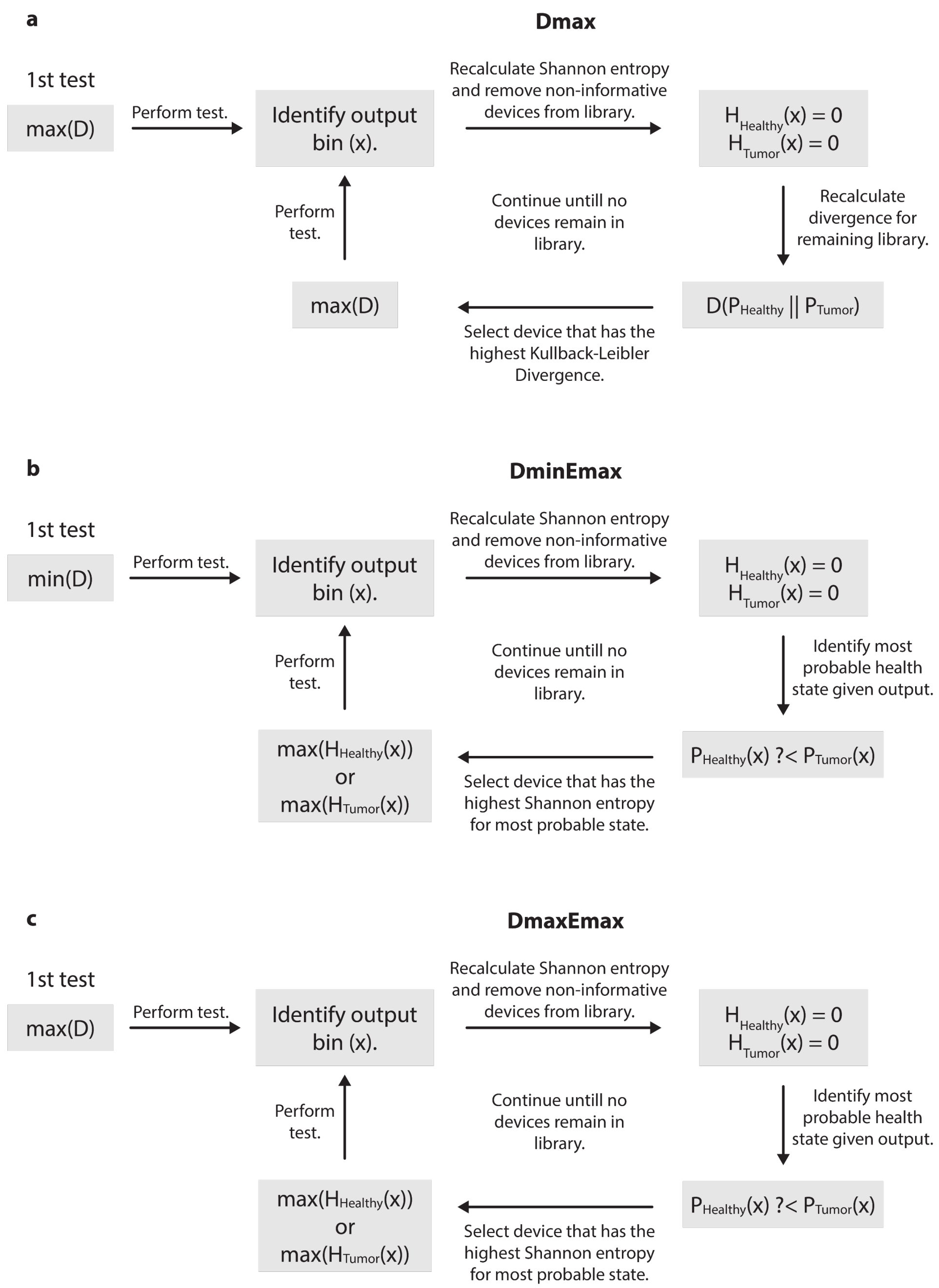

**Supplementary Fig. 16 l Dynamic information theory-based diagnostic strategies.**

**a**, Dmax strategy: First and all subsequent CDAs tested maximize divergence based on the binning output of previous tests.

**b**, DminEmax strategy: First CDA tested has the minimal divergence in library. All subsequent tests use the CDA that has the maximum entropy for the most probable health state.

**c**, DmaxEmax strategy: First CDA tested has the maximum divergence in library. All subsequent tests use the CDA that has the maximum entropy for the most probable health state.
